# Abscess Complications and Prolonged Care in Five-Biomarker-Defined Hypervirulent *Klebsiella pneumoniae* Bloodstream Infection

**DOI:** 10.64898/2026.04.10.26350004

**Authors:** Naoki Watanabe, Tomohisa Watari, Yoshihito Otsuka, Tomoh Matsumiya

**Author notes:** **Corresponding Author:** Dr Naoki Watanabe Department of Bioscience and Laboratory Medicine, Hirosaki University Graduate School of Health Sciences, 66-1 Honcho, Hirosaki, Aomori 036-8564, Japan.

## Abstract

**Background:** Five-biomarker-defined hypervirulent *Klebsiella pneumoniae* (hvKp) causes invasive infections, but its burden in bloodstream infections versus classical *K. pneumoniae* (cKp) is unclear.

**Methods:** This retrospective cohort study at a tertiary hospital in Japan included *K. pneumoniae* bloodstream infection episodes from January 2022–December 2024. hvKp was defined by the presence of all 5 genotypic biomarkers (*rmpA*, *rmpA2*, *iucA*, *iroB*, and *peg-344*). The primary outcome was abscess complications, and secondary outcomes were length of stay and antibiotic duration. Whole-genome sequencing was performed for 164 isolates.

**Results:** Among the 207 episodes, 28 (14%) were of hvKp. Abscess complication occurred in 17 (61%) hvKp versus 23 (13%) cKp episodes (adjusted odds ratio 10.7; 95% CI, 4.36–26.2). Median length of stay in hvKp versus cKp was 28 versus 14 days (adjusted ratio 1.60; 95% CI, 1.18–2.16) and median antibiotic duration was 43 versus 14 days (adjusted ratio 2.13; 95% CI, 1.64–2.77). These associations were attenuated after adjusting for abscess-related complications. No significant difference in 30-day mortality was observed, although the study was underpowered. Multidrug resistance was less frequent in hvKp strains than in cKp strains (11% vs. 30%; P = .040). Among the sequenced hvKp episodes, abscess rates varied across lineages, from 9 of 10 in ST23 to 1 of 4 in ST412.

**Conclusions:** Five biomarker-defined hvKp strains delineated a bloodstream infection subgroup with frequent abscess complications and prolonged care. hvKp and cKp present distinct clinical challenges; diagnostic tools distinguishing these subgroups may aid abscess evaluation and source control.

## Introduction

*Klebsiella pneumoniae* is a leading cause of bloodstream infection and was linked to approximately 790,000 deaths globally in 2019 [1]. Classical *K. pneumoniae* (cKp) predominantly causes infections in patients with comorbidities or immunocompromised. Conversely, hypervirulent *K. pneumoniae* (hvKp) can cause invasive, metastatic infections, including liver abscess, endophthalmitis, and meningitis, even in previously healthy individuals [2]. ST23 is the dominant hvKp lineage, although ST65 and ST86 are also well recognized [3]. In 2024, reports of hvKp acquiring carbapenem-resistance genes triggered alerts from WHO and the European Centre for Disease Prevention and Control, highlighting the need for reliable identification of this pathotype [4,5].

Definitions of hvKp vary across studies. The combination of all five genotypic biomarkers (*iucA*, *iroB*, *peg-344*, *rmpA*, and *rmpA2*) has demonstrated high diagnostic accuracy for identifying hvKp [6–8]. Among *K. pneumoniae* bloodstream infections, the prevalence of hvKp ranges from approximately 4% in the USA and Norway to 26% in Japan [9–11]; nevertheless, hvKp was defined differently across these studies. Consequently, it remains unclear whether hvKp defined by the five-biomarker set is associated with a distinct clinical burden in bloodstream infection and whether this burden differs across lineages.

In this retrospective cohort study at a tertiary hospital in Japan, we aimed to compare the clinical burden of five-biomarker-defined hvKp and cKp bloodstream infection. The primary and secondary outcomes were abscess complication as well as post-onset length of stay, total antibiotic duration, and 30-day mortality, respectively. Moreover, we summarized the genomic features of hvKp isolates, including lineage-specific patterns, in abscess complications.

## METHODS

### Study Design and Data Collection

In this single-center, retrospective cohort study conducted at Kameda Medical Center, a 907-bed tertiary hospital in Kamogawa, Japan, all consecutive patients from whom the *K. pneumoniae* species complex was isolated from at least 1 set of blood cultures between January 1, 2022, and December 31, 2024, were included. Infectious disease consultations were routinely provided for patients with bloodstream infection at this institution. For patients with multiple episodes, only the first episode was included. Of 290 unique episodes identified, two were excluded because of insufficient clinical data or unavailability of the stored isolate (Figure 1). Species-level differentiation by polymerase chain reaction (PCR) [12] identified 81 *K. variicola* or *K. quasipneumoniae* isolates; these were excluded, leaving 207 *K. pneumoniae* isolates for analysis. Clinical and microbiological data were retrospectively collected from electronic medical records and laboratory information systems, and sex was recorded according to the medical records. Blood cultures were processed using the BD BACTEC FX system (Becton Dickinson, Franklin Lakes, NJ, USA), and species identification was performed using a MALDI Biotyper (Bruker Daltonics, Bremen, Germany). Antimicrobial susceptibility testing was performed by the broth microdilution method using a Dry Plate EIKEN (Eiken Chemical, Tokyo, Japan), and results were interpreted according to CLSI M100, 34th edition [13]. The study was approved by the Research Ethics Committee of the Kameda Medical Center (approval number 25-016) with a waiver for informed consent and an opt-out process.

**Figure 1.**
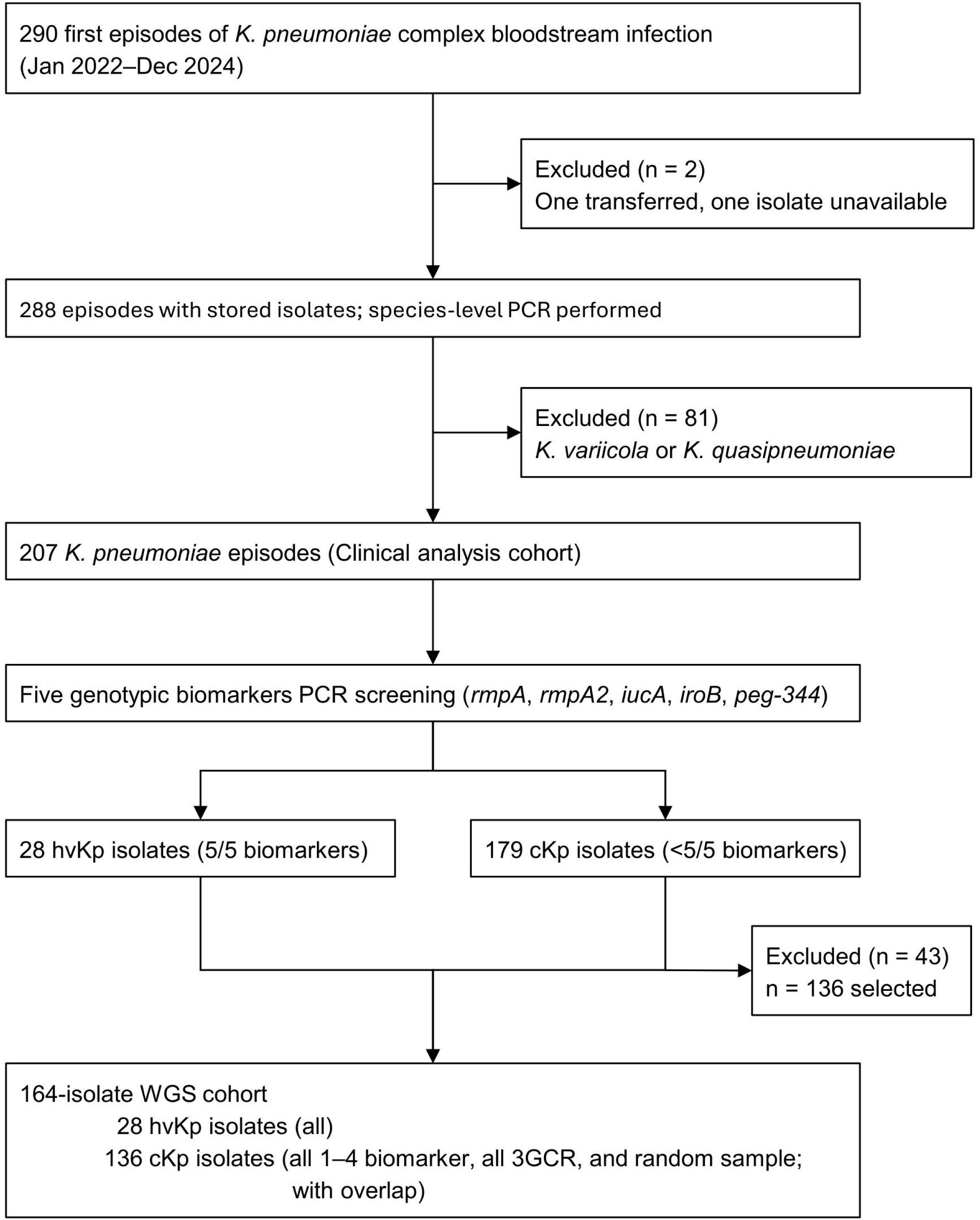
Study Flow Diagram. Flow diagram of episode inclusion in the clinical cohort and isolate selection for whole-genome sequencing. Of the 290 first episodes of *Klebsiella pneumoniae* complex bloodstream infection, 207 *K. pneumoniae* episodes were included in the clinical analysis. Whole-genome sequencing was performed on 164 isolates using an enriched sampling strategy comprising all hvKp isolates, all isolates with 1–4 biomarkers, all 3GCR isolates, and a random sample of the remaining cKp isolates with overlap between categories. Abbreviations: 3GCR, third-generation cephalosporin-resistant; cKp, classical *Klebsiella pneumoniae*; hvKp, hypervirulent *Klebsiella pneumoniae*; PCR, polymerase chain reaction; WGS, whole-genome sequencing.

### Definitions

hvKp was defined by the presence of all five genotypic biomarkers (*iucA*, *iroB*, *peg-344*, *rmpA*, and *rmpA2*), as proposed by Russo et al. [6–8] (Supplementary Methods). All other isolates were classified as cKp. Screening was performed sequentially: *rmpA* was tested first, and the remaining markers were tested only in *rmpA*-positive isolates. The hvKp classification was assigned retrospectively for study analyses and was not available to the treating clinicians during routine care. Third-generation cephalosporin resistance (3GCR) was defined as nonsusceptibility to ceftriaxone. Multidrug resistance was defined as nonsusceptibility to at least one agent in three or more antimicrobial categories [14]. Twelve of the 15 antimicrobial categories specified for *Enterobacteriaceae* in this framework were tested; glycylcyclines, phosphonic acids, and polymyxins were not routinely tested at our institution and were excluded from the classification. Infection onset was classified as community-acquired if the index blood culture was collected within 48 h of admission without prior healthcare exposure. All other episodes were classified as healthcare-associated. The Charlson Comorbidity Index was calculated after excluding age points [15]. Abscess complications were defined based only on imaging findings, and clinically suspected abscesses without imaging confirmation were excluded. Metastatic infection was defined as a secondary infectious focus anatomically distinct from the presumed primary source of bloodstream infection, including abscesses and non-suppurative manifestations such as endophthalmitis. The post-onset length of stay (LOS) was defined as the number of days from index blood culture collection to hospital discharge or transfer. Total antibiotic duration was determined as the total number of days of antimicrobial therapy for a bloodstream infection episode and its complications, including oral therapy after discharge. Detailed operational definitions are provided in the Supplementary Methods.

### Whole-Genome Sequencing

Whole-genome sequencing (WGS) was performed on a subset of isolates selected to maximize representation of biomarker-positive and antimicrobial-resistant isolates within the available sequencing budget. The sequenced subset comprised all hvKp isolates, all isolates with 1–4 of the five positive markers, all 3GCR- or carbapenem-resistant isolates, and a random sample of the remaining cKp isolates, with overlap between groups. *De novo* assembly was performed using Unicycler [16]. Assembly quality was assessed using QUAST and CheckM2 with predefined quality criteria [17,18]. Genomic characterization encompassed multilocus sequence typing, capsular locus typing, virulence gene profiling, and resistance gene detection, performed using Pathogenwatch, Kleborate, and AMRFinderPlus [19–21]. A recombination-filtered core genome single-nucleotide polymorphism (SNP) phylogeny was inferred by mapping reads to the *K. pneumoniae* NTUH-K2044 reference genome using Snippy, removing recombinant regions with gubbins, and constructing a maximum-likelihood tree with IQ-TREE [22–24]. For hvKp isolates carrying ESBL (extended-spectrum β-lactamase) genes, contig-level analysis was performed to assess the genomic context of resistance and virulence determinants. Additional technical details are provided in the Supplementary Methods.

### Statistical Analysis

Statistical analyses were performed using EZR version 1.70 [25]. Continuous variables were compared using the Mann-Whitney U test, and categorical variables were compared using Fisher’s exact test. All tests were two-sided, with statistical significance set at p<.05. The primary outcome was abscess complications; secondary outcomes were post-onset length of stay and total antibiotic duration, and 30-day mortality was evaluated as a supportive outcome.

Covariates were selected a priori based on clinical relevance as baseline confounders, and the number of covariates was constrained by the number of outcome events to avoid overfitting. Abscess complications were evaluated using multivariable logistic regression adjusted for age, sex, and the Charlson Comorbidity Index. For 30-day mortality, Firth’s penalized logistic regression was used because the number of death events was limited relative to the number of candidate covariates [26]. The adjustment set comprised age and the Charlson Comorbidity Index. For secondary outcomes, two sequential models were fitted: Model A estimated the overall association of hvKp with each outcome, and Model B additionally included abscess complications to assess attenuation. Length of stay was analyzed as ln(LOS+1) using multivariable linear regression, and coefficients were exponentiated to report adjusted ratios with 95% confidence intervals. Total antibiotic duration was analyzed among the treatment completers. Antibiotic duration was analyzed after log transformation using multivariable linear regression. A common base adjustment set comprising age, sex, Charlson Comorbidity Index, and infection acquisition category was used for both models. For antibiotic duration, the primary infection source was also included because treatment duration was strongly influenced by the infectious syndrome and source-control requirements. For each outcome, episodes with missing data were excluded from the corresponding analysis. Sensitivity and threshold analyses are described in the Supplementary Methods.

As a prespecified exploratory analysis, outcomes were stratified by biomarker count (0, 1–4, or 5 of 5); the 1–4-marker group was presented descriptively because of the small sample size. Genomic features of the hvKp isolates, including lineage, capsule type, and virulence plasmid structure, were summarized descriptively rather than formally compared across lineages.

## RESULTS

### Patient Characteristics

Among 207 episodes of *K. pneumoniae* bloodstream infection, 28 (14%; 95% confidence interval [CI] 9–19) were classified as hvKp and 179 (86%; 95% CI 81–91) as cKp (Table 1 and Table S1). Median age was similar between groups (76 vs. 78 years), and the proportion of male patients did not differ significantly (75% vs. 60%). Patients with hvKp had fewer comorbidities; the proportion with a Charlson Comorbidity Index score ≥2 was lower in the hvKp group than in the cKp group (57% vs. 80%). Markers of healthcare exposure, including indwelling devices (43% vs. 64%) and prior antibiotic use (32% vs. 55%), were also less frequent. Primary infection sources were broadly similar between the groups, although respiratory infection and unknown source were more frequent in the hvKp and cKp, respectively. Respiratory infection remained uncommon overall, accounting for 9 of 207 episodes (4 %). Multidrug resistance was less common in hvKp than in cKp (11% vs. 30%; Table S2). Appropriate empiric antimicrobial therapy was administered in 27 of 28 hvKp episodes (96%) and 151 of 179 cKp episodes (84%). The missing and analyzed populations for each outcome are summarized in Table S3.

**Table 1.**
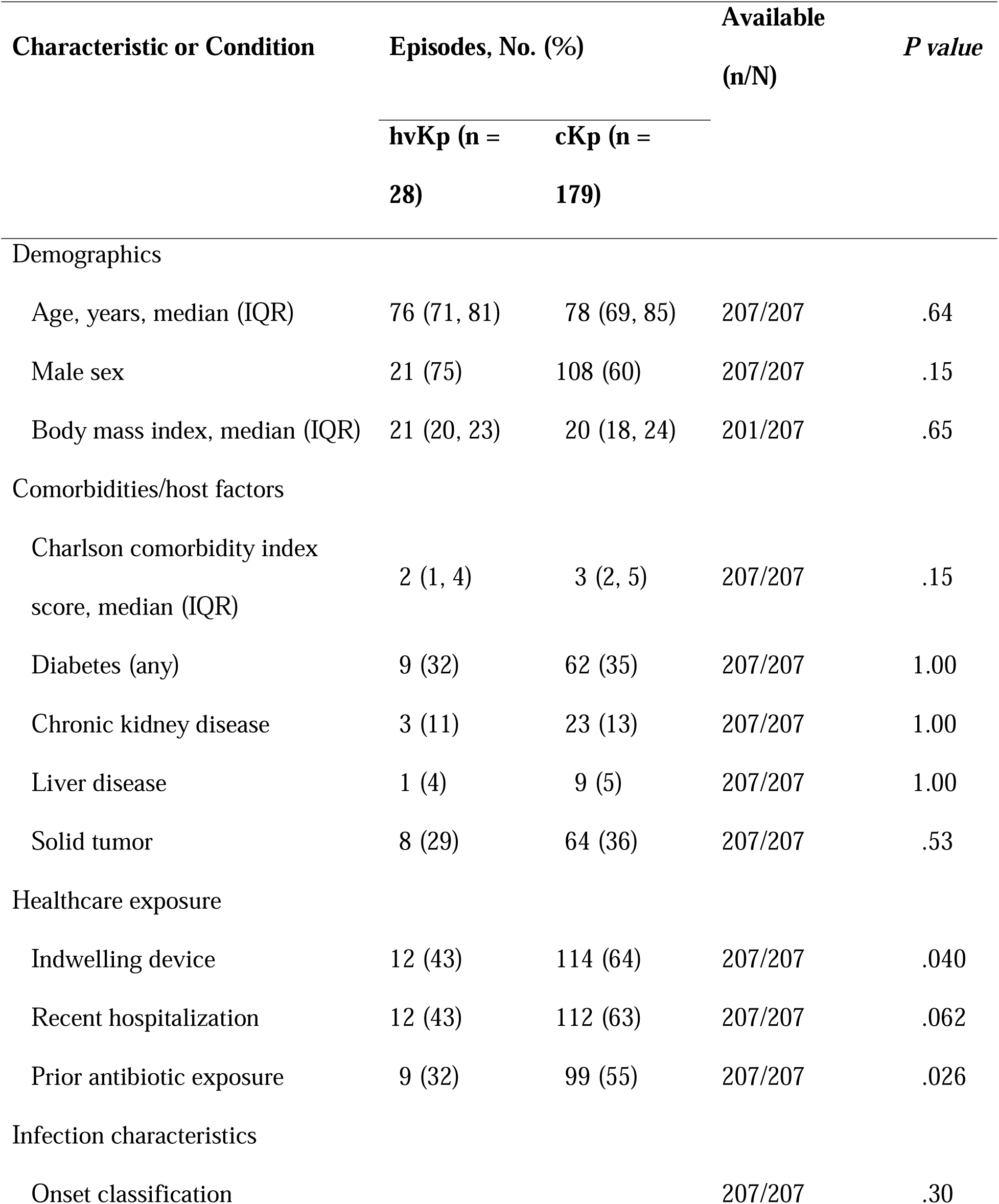
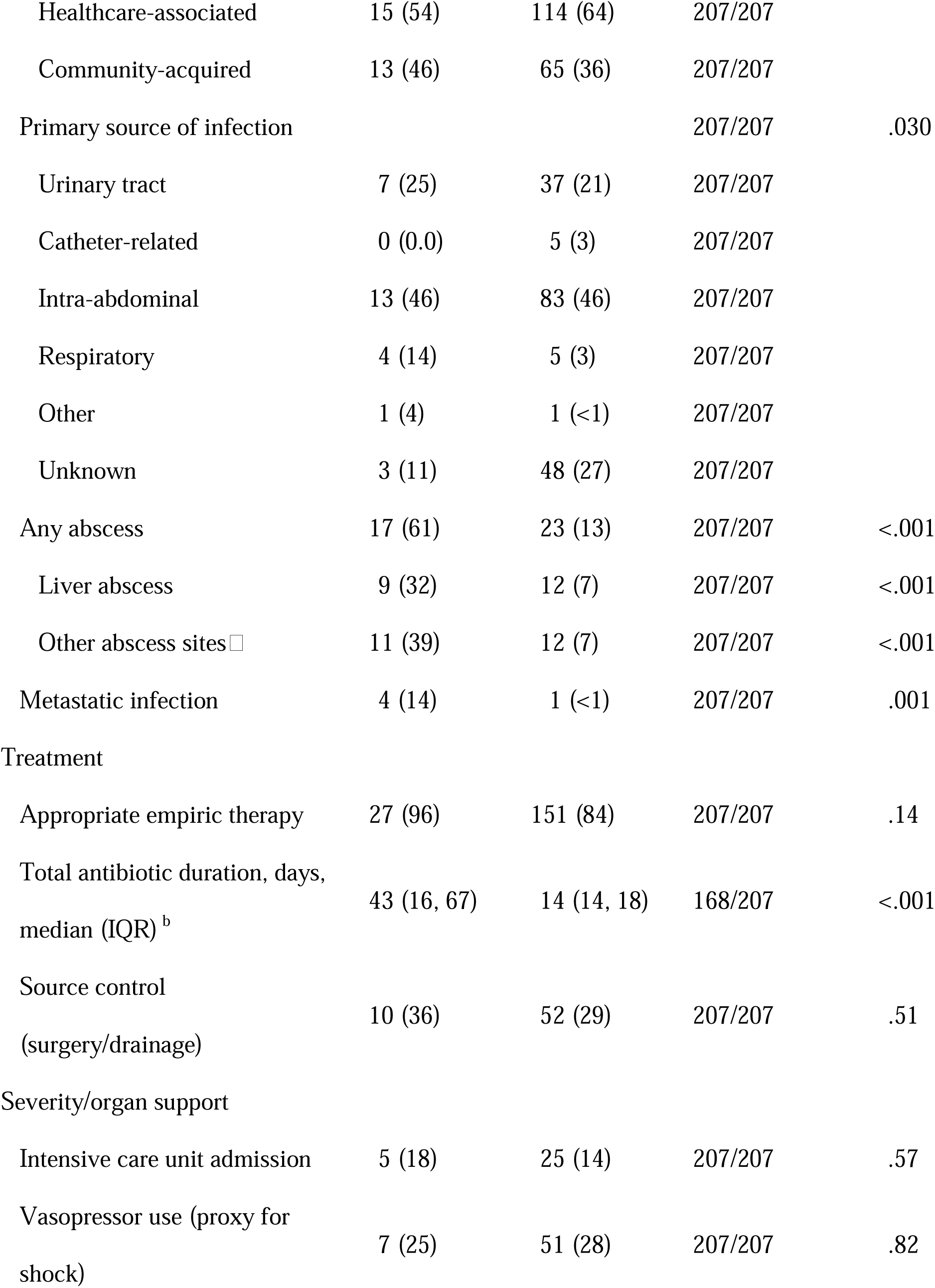
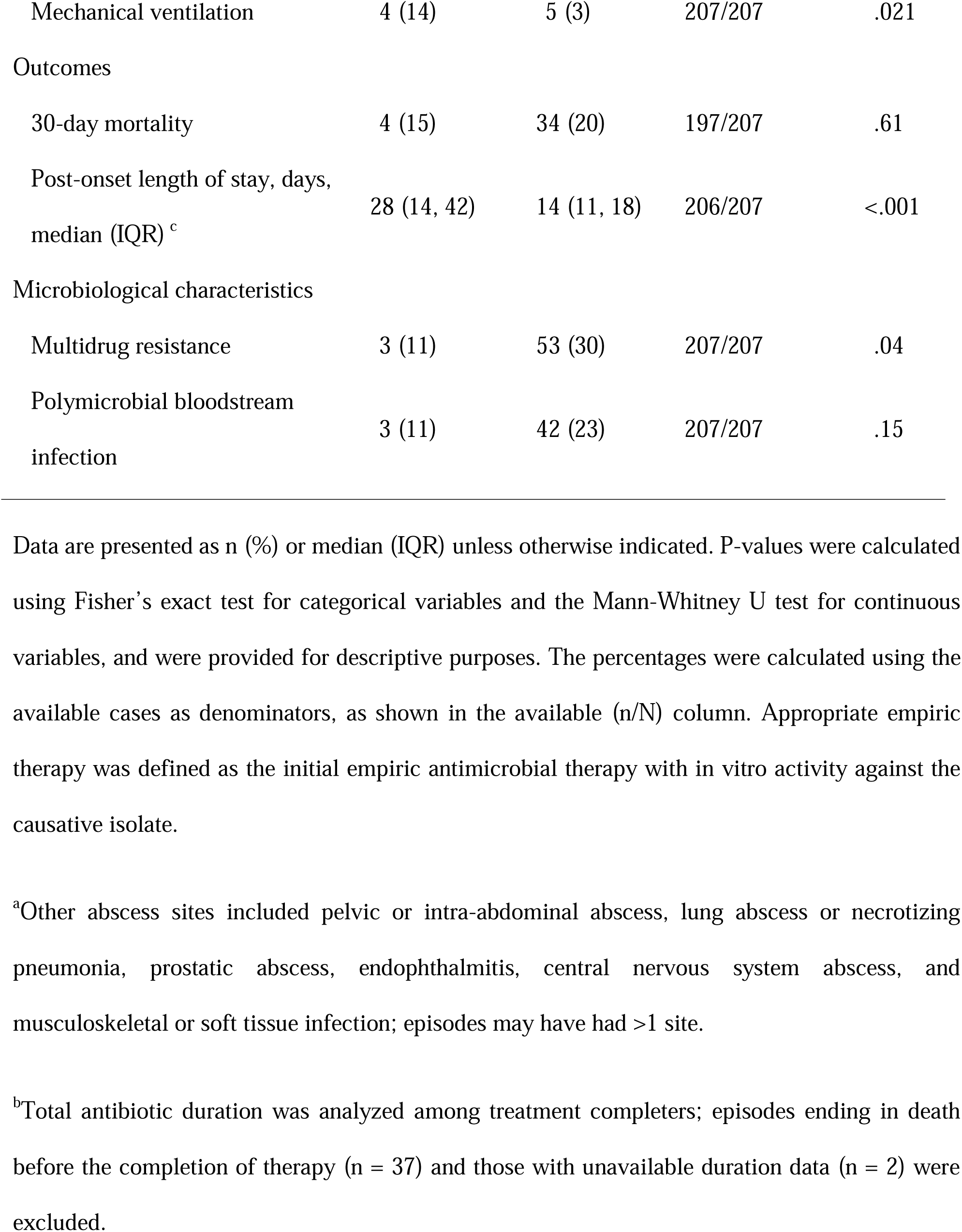
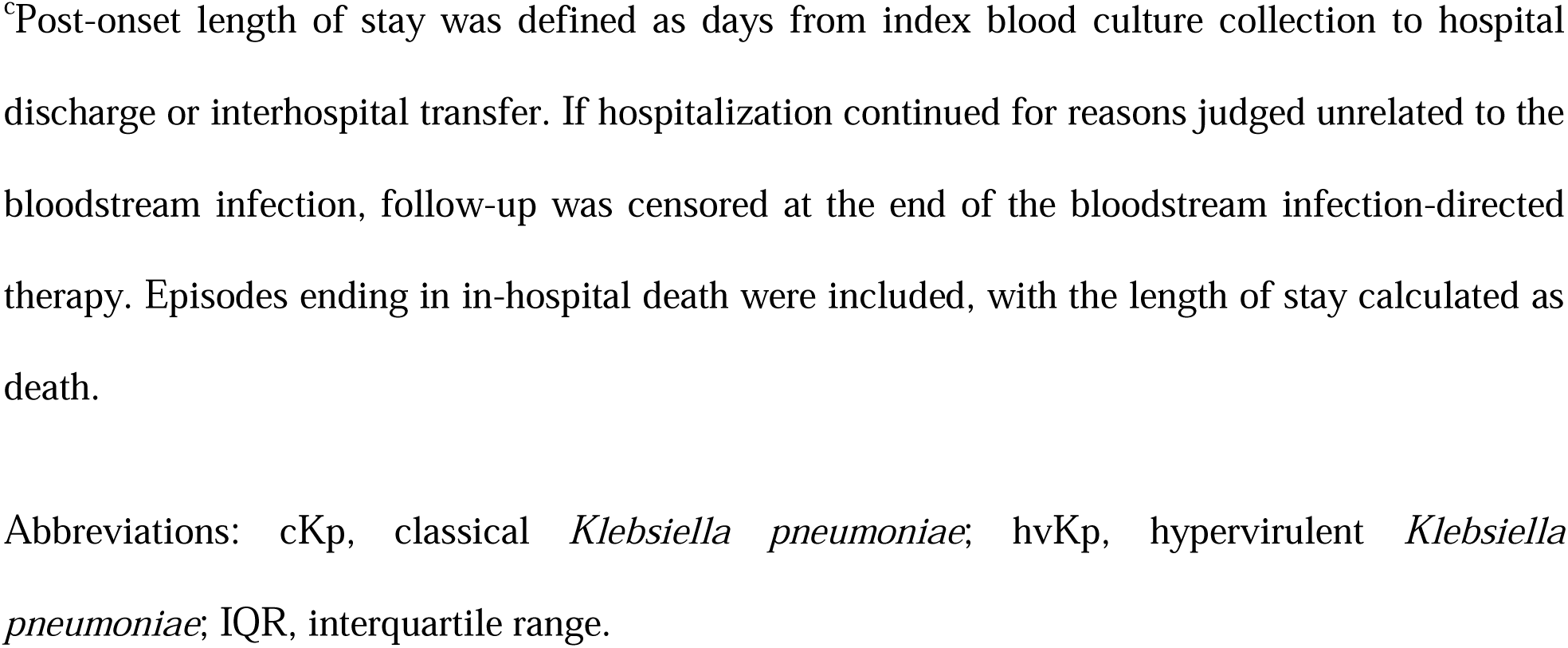
Characteristics and Clinical Outcomes of *Klebsiella pneumoniae* Bloodstream Infection Episodes, by Hypervirulent Status. Data are presented as n (%) or median (IQR) unless otherwise indicated. P-values were calculated using Fisher’s exact test for categorical variables and the Mann-Whitney U test for continuous variables, and were provided for descriptive purposes. The percentages were calculated using the available cases as denominators, as shown in the available (n/N) column. Appropriate empiric therapy was defined as the initial empiric antimicrobial therapy with in vitro activity against the causative isolate. ^a^Other abscess sites included pelvic or intra-abdominal abscess, lung abscess or necrotizing pneumonia, prostatic abscess, endophthalmitis, central nervous system abscess, and musculoskeletal or soft tissue infection; episodes may have had >1 site. ^b^Total antibiotic duration was analyzed among treatment completers; episodes ending in death before the completion of therapy (n = 37) and those with unavailable duration data (n = 2) were excluded. ^c^Post-onset length of stay was defined as days from index blood culture collection to hospital discharge or interhospital transfer. If hospitalization continued for reasons judged unrelated to the bloodstream infection, follow-up was censored at the end of the bloodstream infection-directed therapy. Episodes ending in in-hospital death were included, with the length of stay calculated as death. Abbreviations: cKp, classical *Klebsiella pneumoniae*; hvKp, hypervirulent *Klebsiella pneumoniae*; IQR, interquartile range.

| Characteristic or Condition | Episodes, No. (%) |  | Available<br>(n/N) | *P value* |
| --- | --- | --- | --- | --- |
|  | hvKp (n = | cKp (n = |  |  |
|  | 28) | 179) |  |  |
| Demographics |  |  |  |  |
| Age, years, median (IQR) | 76 (71, 81) | 78 (69, 85) | 207/207 | .64 |
| Male sex | 21 (75) | 108 (60) | 207/207 | .15 |
| Body mass index, median (IQR) | 21 (20, 23) | 20 (18, 24) | 201/207 | .65 |
| Comorbidities/host factors |  |  |  |  |
| Charlson comorbidity index score, median (IQR) | 2 (1, 4) | 3 (2, 5) | 207/207 | .15 |
| Diabetes (any) | 9 (32) | 62 (35) | 207/207 | 1.00 |
| Chronic kidney disease | 3 (11) | 23 (13) | 207/207 | 1.00 |
| Liver disease | 1 (4) | 9 (5) | 207/207 | 1.00 |
| Solid tumor | 8 (29) | 64 (36) | 207/207 | .53 |
| Healthcare exposure |  |  |  |  |
| Indwelling device | 12 (43) | 114 (64) | 207/207 | .040 |
| Recent hospitalization | 12 (43) | 112 (63) | 207/207 | .062 |
| Prior antibiotic exposure | 9 (32) | 99 (55) | 207/207 | .026 |
| Infection characteristics |  |  |  |  |
| Onset classification |  |  | 207/207 | .30 |
| Healthcare-associated | 15 (54) | 114 (64) | 207/207 |  |
| Community-acquired | 13 (46) | 65 (36) | 207/207 |  |
| Primary source of infection |  |  | 207/207 | .030 |
| Urinary tract | 7 (25) | 37 (21) | 207/207 |  |
| Catheter-related | 0 (0.0) | 5 (3) | 207/207 |  |
| Intra-abdominal | 13 (46) | 83 (46) | 207/207 |  |
| Respiratory | 4 (14) | 5 (3) | 207/207 |  |
| Other | 1 (4) | 1 (<1) | 207/207 |  |
| Unknown | 3 (11) | 48 (27) | 207/207 |  |
| Any abscess | 17 (61) | 23 (13) | 207/207 | <.001 |
| Liver abscess | 9 (32) | 12 (7) | 207/207 | <.001 |
| Other abscess sites<sup>□</sup> | 11 (39) | 12 (7) | 207/207 | <.001 |
| Metastatic infection | 4 (14) | 1 (<1) | 207/207 | .001 |
| Treatment |  |  |  |  |
| Appropriate empiric therapy | 27 (96) | 151 (84) | 207/207 | .14 |
| Total antibiotic duration, days,<br>median (IQR)<sup>b</sup> | 43 (16, 67) | 14 (14, 18) | 168/207 | <.001 |
| Source control<br>(surgery/drainage) | 10 (36) | 52 (29) | 207/207 | .51 |
| Severity/organ support |  |  |  |  |
| Intensive care unit admission | 5 (18) | 25 (14) | 207/207 | .57 |
| Vasopressor use (proxy for<br>shock) | 7 (25) | 51 (28) | 207/207 | .82 |
| Mechanical ventilation | 4 (14) | 5 (3) | 207/207 | .021 |
| Outcomes |  |  |  |  |
| 30-day mortality | 4 (15) | 34 (20) | 197/207 | .61 |
| Post-onset length of stay, days,<br>median (IQR) <sup>c</sup> | 28 (14, 42) | 14 (11, 18) | 206/207 | <.001 |
| Microbiological characteristics |  |  |  |  |
| Multidrug resistance | 3 (11) | 53 (30) | 207/207 | .04 |
| Polymicrobial bloodstream<br>infection | 3 (11) | 42 (23) | 207/207 | .15 |

**Table 2.**
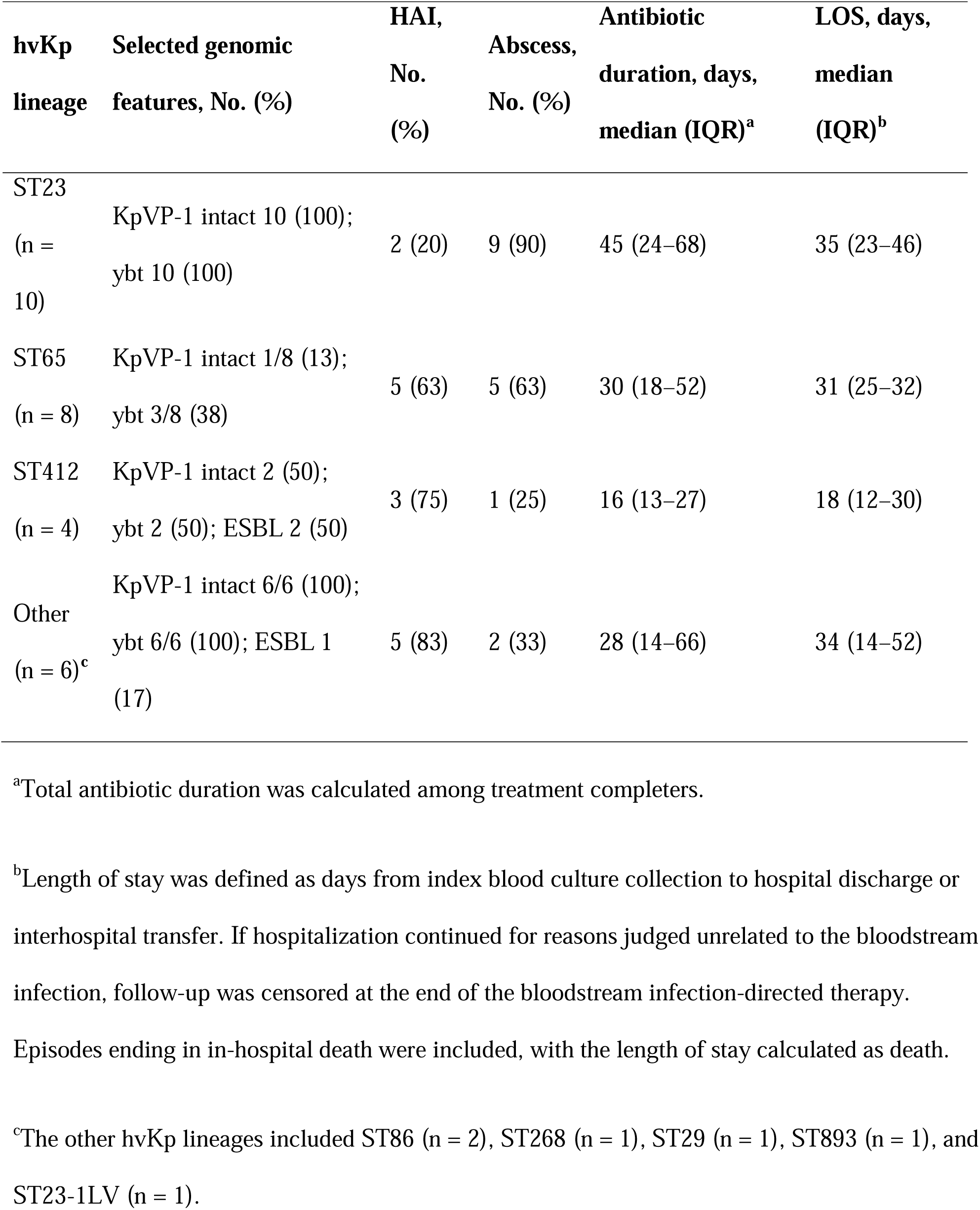
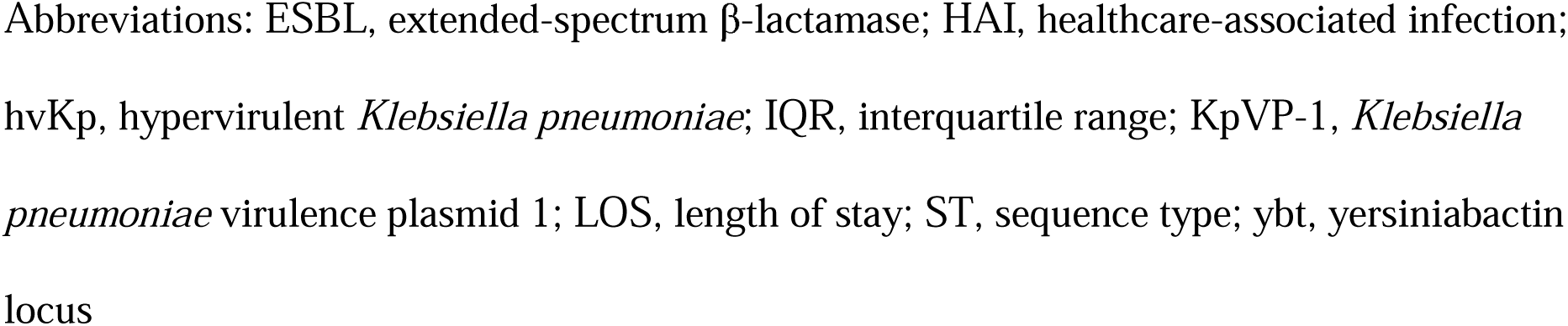
Lineage-Specific Genomic and Clinical Features of Hypervirulent *Klebsiella pneumoniae* Bloodstream Isolates. ^a^Total antibiotic duration was calculated among treatment completers. ^b^Length of stay was defined as days from index blood culture collection to hospital discharge or interhospital transfer. If hospitalization continued for reasons judged unrelated to the bloodstream infection, follow-up was censored at the end of the bloodstream infection-directed therapy. Episodes ending in in-hospital death were included, with the length of stay calculated as death. ^c^The other hvKp lineages included ST86 (n = 2), ST268 (n = 1), ST29 (n = 1), ST893 (n = 1), and ST23-1LV (n = 1). Abbreviations: ESBL, extended-spectrum β-lactamase; HAI, healthcare-associated infection; hvKp, hypervirulent *Klebsiella pneumoniae*; IQR, interquartile range; KpVP-1, *Klebsiella pneumoniae* virulence plasmid 1; LOS, length of stay; ST, sequence type; ybt, yersiniabactin locus

| hvKp lineage | Selected genomic features, No. (%) | HAI, No. (%) | Abscess, No. (%) | Antibiotic duration, days, median (IQR)<sup>a</sup> | LOS, days, median (IQR)<sup>b</sup> |
| --- | --- | --- | --- | --- | --- |
| ST23<br>(n = 10) | KpVP-1 intact 10 (100);<br>ybt 10 (100) | 2 (20) | 9 (90) | 45 (24–68) | 35 (23–46) |
| ST65<br>(n = 8) | KpVP-1 intact 1/8 (13);<br>ybt 3/8 (38) | 5 (63) | 5 (63) | 30 (18–52) | 31 (25–32) |
| ST412<br>(n = 4) | KpVP-1 intact 2 (50);<br>ybt 2 (50); ESBL 2 (50) | 3 (75) | 1 (25) | 16 (13–27) | 18 (12–30) |
| Other<br>(n = 6)<sup>c</sup> | KpVP-1 intact 6/6 (100);<br>ybt 6/6 (100); ESBL 1<br>(17) | 5 (83) | 2 (33) | 28 (14–66) | 34 (14–52) |

### Abscess Complications

Abscess complications occurred in 17 of 28 hvKp episodes (61%; 95% CI 41–79) versus 23 of 179 cKp episodes (13%; 95% CI 8–19; p<0.001). In multivariable logistic regression, adjusted for age, sex, and Charlson comorbidity index, hvKp was independently associated with abscess complications (adjusted odds ratio [OR] 10.7; 95% CI 4.36–26.2; p<0.001; Table S4). Liver abscess was the most common hvKp site (32% vs. 7%), and abscesses at other sites were also more frequent (39% vs. 7%); some patients had multiple sites. Extrahepatic sites included urogenital, intra-abdominal, central nervous system, pulmonary, musculoskeletal, and ocular locations, some of which occurred exclusively in hvKp (Table S1). Metastatic infection was identified in 4 of 28 hvKp episodes (14%) compared with 1 of 179 cKp episodes (<1%; p=0.001). Abscess complication rates rose with the number of biomarkers, occurring in 12%, 25%, and 61% of the 0-marker, 1–4-marker, and 5-marker groups, respectively (Table S5).

### Length of Stay, Antibiotic Duration, and 30-Day Mortality

The post-onset length of stay data were available for 206 episodes (28 hvKp and 178 cKp). The length of stay was longer for hvKp than for cKp (median 28 days [interquartile range 14–42] vs. 14 days [11–18]; *p* < 0.001). For the antibiotic duration, 39 episodes were excluded, primarily due to death before treatment completion. Among the remaining 168 treatment completers (24 hvKp and 144 cKp), the total antibiotic duration was similarly longer (43 days [16–67] vs. 14 days [14–18]; p<.001). In adjusted analyses, hvKp was associated with longer length of stay (adjusted ratio 1.60; 95% CI 1.18–2.16) and antibiotic duration (adjusted ratio 2.13; 95% CI 1.64–2.77) (Tables S6 and S7). These associations were attenuated after additional adjustment for abscess complication (adjusted ratio 1.07; 95% CI 0.79–1.45 for length of stay; 1.24; 1.00–1.54 for antibiotic duration). Abscess complication remained associated with prolonged hospitalization (2.35, 1.80–3.06) and prolonged therapy (3.08, 2.53–3.76; Figure 2). The prespecified threshold analyses of prolonged antibiotic duration aligned with these findings (Table S8).

**Figure 2.**
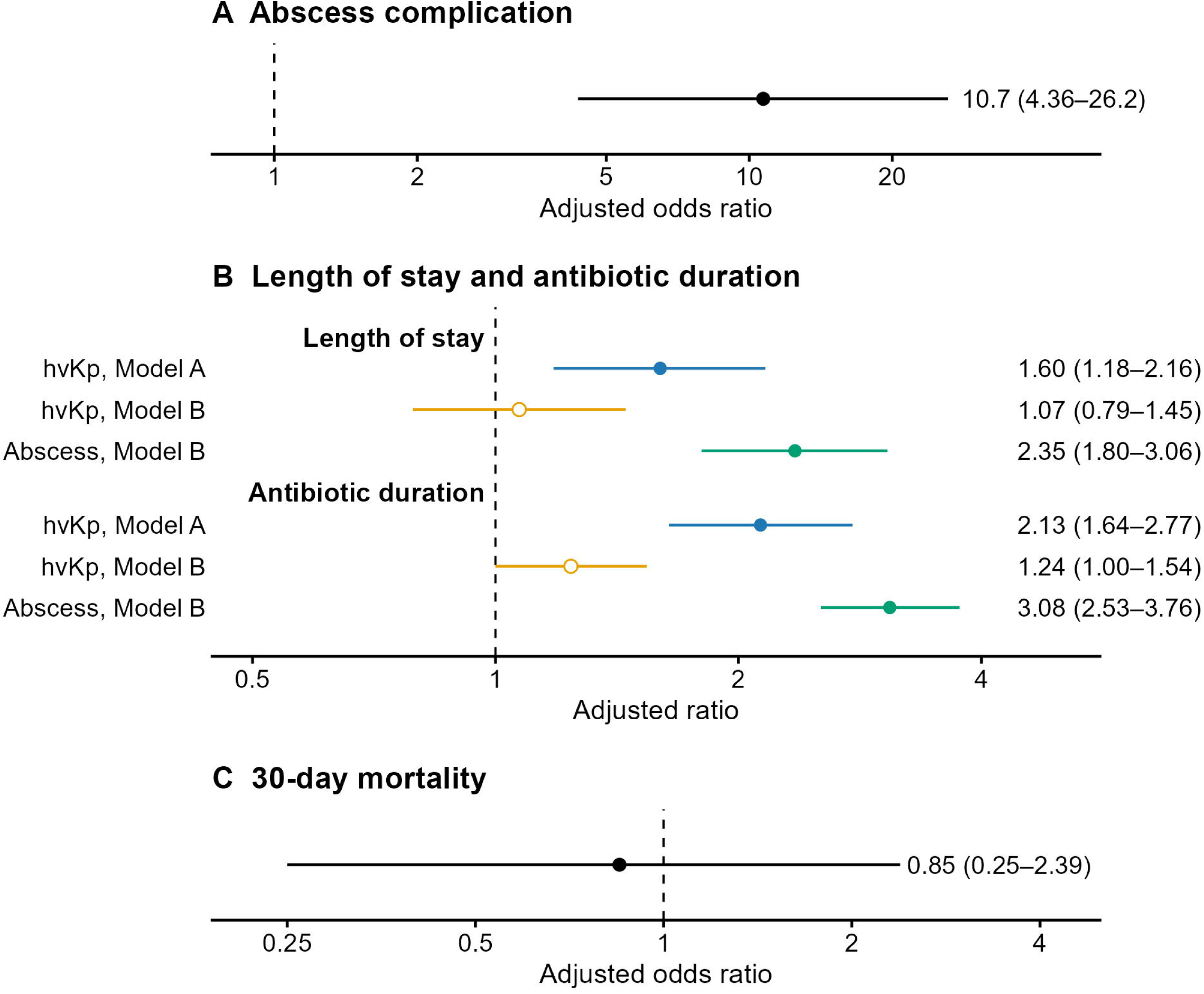
Association of Hypervirulent *K. pneumoniae* With Clinical Outcomes. (A) Adjusted odds ratio for abscess complications. (B) Adjusted ratios for log-transformed length of stay and total antibiotic duration among treatment completers. In panel B, Model A presents the association of hvKp without abscess complications, and Model B shows the association of hvKp after additional adjustment for abscess complications. (C) Adjusted odds ratio for 30-day mortality. Circles indicate point estimates and horizontal lines indicate 95% confidence intervals. The dashed vertical line indicates a null value of 1.0. The x-axis scales differ between the panels. Abscess complications were assessed in 207 episodes; length of stay in 206 episodes with available data; antibiotic duration in 168 treatment completers; and 30-day mortality in 197 episodes with available vital statuses. The full model specifications are listed in Supplementary Tables S4, S6, S7, and S9. Abbreviations: cKp, classical *Klebsiella pneumoniae*; hvKp, hypervirulent *Klebsiella pneumoniae*.

Thirty-day mortality was 15% (4/27; 95% CI 4–34) and 20% (34/170; 95% CI 14–27) in hvKp and cKp, respectively (p=0.61); episodes with unknown vital status were excluded. In adjusted analyses, the OR for 30-day mortality associated with hvKp was 0.85 (95% CI 0.25–2.39; p=0.77; Table S9). Intensive care unit admission rates were 18% and 14% in the hvKp and cKp groups, respectively, and vasopressor use was 25% and 29%, respectively. Mechanical ventilation was more frequent in the hvKp group (14% vs. 3%); nonetheless, adjusted analysis was not performed because of the small number of events.

### Genomic Characteristics and Lineage-Specific Findings

WGS was performed on 164 isolates, including 28 hvKp and 136 non-hvKp isolates. The non-hvKp group comprised a random sample of 125 cKp isolates, with additional selection to ensure complete representation of isolates with markers 1–4 (n = 8) and 3GCR (n = 31). No carbapenem-resistant isolates were identified among the 207 episodes. All assemblies met predefined quality criteria. The clinical characteristics were similar between the randomly sampled sequenced cKp isolates (n = 125) and non-sequenced cKp isolates (n = 54), except for the distribution of primary infection sources (Table S10).

Sequence type and capsular (KL) locus distributions are provided in Tables S11 and S12, respectively. Among hvKp isolates (n = 28), a limited number of lineages predominated, most commonly ST23/KL1 and ST65/KL2. ST23/KL1 and ST65/KL2 accounted for 10 (36%) and eight (29%) isolates, respectively, followed by ST412/KL57 and ST86/KL2 (n = 4, 14% and n = 2, 7%, respectively). Conversely, the randomly sampled cKp isolates (n = 125) exhibited substantial sequence type diversity, comprising 75 sequence types; ST37 and ST45 were the most common (n = 10, 8% and n = 9, 7%, respectively). Among the 3GCR isolates (n = 31), three lineages predominated: ST307/KL102, ST37/KL15, and ST353/KL110 (n = 8, 26%; n = 7, 23%; and n = 7, 23%, respectively).

The recombination-filtered core-genome SNP phylogeny indicated that hvKp isolates were distributed across multiple lineages rather than clustering in a single clade (Figure 3). Abscess complication rates among hvKp episodes differed by lineage (Table S13): 9/10 (90%) in ST23/KL1, 5/8 (63%) in ST65/KL2, 1/4 (25%) in ST412/KL57, and 2/6 (33%) in other lineages. Genomic differences were also observed across hvKp lineages. ST23/KL1 isolates uniformly carried intact KpVP-1 and yersiniabactin, whereas ST65/KL2 isolates more frequently had KpVP-1 truncation (7/8).

**Figure 3.**
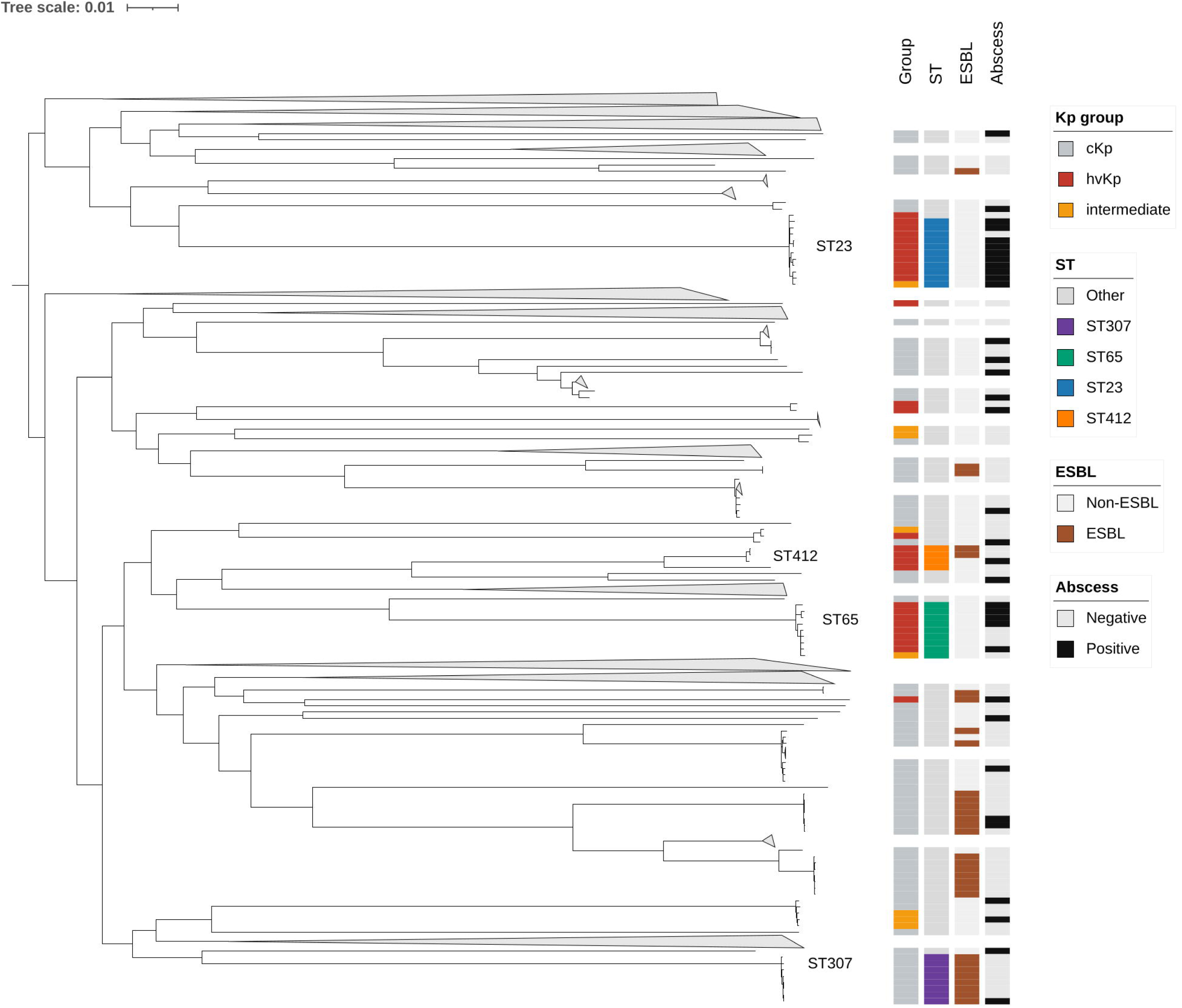
Phylogenetic Structure of Bloodstream *K. pneumoniae* Isolates and Lineage-Specific Distribution of Hypervirulence, ESBL Status, and Abscess Complication. Maximum likelihood phylogeny of 164 bloodstream *Klebsiella pneumoniae* isolates from Japan inferred from a recombination-filtered core genome single-nucleotide polymorphism alignment. Annotation tracks indicate the study group (hvKp, intermediate [1–4 biomarkers], or cKp), selected sequence types (ST23, ST65, ST307, ST412, or others), ESBL status, and abscess complications. Major clades were labeled directly on the tree, and non-focal cKp lineages were collapsed for clarity. The annotation is intended to illustrate the distribution of hypervirulence, resistance, and abscess complications across phylogenies. Lineage-specific comparisons are descriptive owing to small subgroup sizes. Abbreviations: cKp, classical *Klebsiella pneumoniae*; ESBL, extended-spectrum β-lactamase; hvKp, hypervirulent *Klebsiella pneumoniae*; ST, sequence type.

hvKp-ESBL convergence was uncommon, accounting for three of 207 episodes (1.5%). Two episodes involved ST412/KL57 isolates harboring *bla*CTX-M-15; these belonged to the same clonal group (CG10014) and were isolated in successive years. The third episode involved an ST893/KL20 isolate carrying *bla*CTX-M-55. All three convergence episodes were healthcare-associated, and none resulted in death. In all three isolates, the *ESBL* gene was located on a contig assigned to a putative plasmid cluster, separate from the contigs carrying virulence-associated sequences (Supplementary Note).

## DISCUSSION

In this retrospective cohort of *K. pneumoniae* bloodstream infections, five-biomarker-defined hvKp isolates were used to stratify subgroups with frequent abscess complications and prolonged care. Associations of hospital length of stay and antibiotic duration with hvKp status were attenuated after additional adjustment for abscess complications. Whole-genome sequencing revealed substantial genomic heterogeneity within hvKp, with lineage-specific differences in abscess frequency.

The prevalence of hvKp in this cohort (14%) aligned with reports from Japan [11] but was higher than that reported in the US and Norway [10,27]. These differences likely reflect both the epidemiological setting and variability in hvKp definitions across studies. Broader definitions based on a subset of biomarkers may classify isolates in the 1–4-marker group as hvKp, thereby yielding higher prevalence estimates than the five-marker definition used here. In this cohort, abscess complication rates rose across biomarker strata, from 12% in the 0-marker group to 25% in the 1–4-marker group and 61% in the five-marker group. This gradient aligns with studies reporting that requiring all five markers provides greater accuracy for identifying hvKp than lower thresholds [7]. Therefore, isolates positive for 1–4 markers may be better considered an intermediate group rather than definitively hvKp. The clinical relevance of this definition is further supported by the distinct burden observed in the five-marker group: frequent abscesses, prolonged hospitalization, and extended treatment.

hvKp bloodstream infections defined by five biomarkers were associated with frequent abscess complications. These findings suggest that the greater care burden associated with hvKp is partly related to abscess-prone presentations. 30-day mortality rates did not differ between the hvKp and cKp groups in our cohort. Nonetheless, this result should be interpreted with caution. Tang et al. reported increased early mortality in genomically defined hvKp cases in a cohort that included diverse infection types [28]. In our cohort, respiratory infections, previously associated with poor outcomes in *K. pneumoniae* bloodstream infection [9], accounted for only 4% of episodes. Therefore, differences in infection-type composition and limited power may have contributed to the discrepant mortality findings.

The clinical challenges posed by *K. pneumoniae* bloodstream infections vary with the prevalence of hvKp. In a US cohort in which hvKp is rare, resistance to the initial prescribed antibiotic was the strongest predictor of mortality [9]. Here, antimicrobial resistance remained an important concern for cKp, which accounted for 86% of episodes and had a higher multidrug-resistance rate than hvKp. Conversely, hvKp isolates were largely susceptible to standard empiric agents, and multidrug resistance was less frequent than in cKp isolates. Collectively, these findings suggest that management priorities differ between subgroups: cKp requires attention to antimicrobial resistance, whereas hvKp requires early abscess evaluation and source control. Therefore, early distinction between hvKp and cKp may aid abscess evaluation and source control.

Beyond the immediate clinical burden of hvKp bloodstream infection, clinically accessible diagnostics and ongoing genomic surveillance remain translational priorities. At present, the five-biomarker PCR for hvKp is a research tool, and no commercial assay is available for routine clinical use [6,8]. Although such assays could be developed on existing PCR platforms [29], they have not yet been validated in clinical laboratories [8]. Distinguishing hvKp from cKp may facilitate abscess evaluation and source control. Another concern is the genomic convergence of hypervirulence and antimicrobial resistance [4]. In this cohort, such convergence was uncommon, accounting for only three of the 207 episodes (1.5%). In all three isolates, ESBL genes were carried on plasmid contigs separate from the virulence-associated sequences, suggesting independent acquisition. Collectively, these findings support the need for clinically accessible hvKp diagnostics and continued genomic surveillance to monitor convergence of hypervirulence and antimicrobial resistance.

The consecutive case design in a tertiary care setting with routine infectious diseases consultation may have enabled characterization of the clinical burden of abscesses in this cohort. Nonetheless, this study had some limitations. First, this retrospective study at a single tertiary hospital may limit the generalizability of our findings. Second, abscess imaging was not protocolized, and imaging-frequency data were unavailable. Because hvKp episodes may more often present with features prompting cross-sectional imaging, differential ascertainment of abscess complications cannot be excluded, which could have contributed to the observed between-group differences in abscess rates. Nevertheless, the large effect size and descriptive gradient in abscess frequency across biomarker strata suggest that ascertainment bias alone is unlikely to fully account for the observed association. Third, the number of hvKp episodes was small, limiting the statistical power of some comparisons, including 30-day mortality. Fourth, whole-genome sequencing was performed on 164 of the 207 isolates. Clinical characteristics were similar between sequenced and non-sequenced cKp isolates, but residual selection bias could not be ruled out.

In conclusion, hvKp, defined by the five-biomarker panel, identifies a bloodstream infection subset characterized by frequent abscess complications and a prolonged clinical course. Distinguishing hvKp from cKp at the time of bloodstream infection may aid abscess evaluation and guide timely source control, reflecting the distinct clinical challenges of each pathotype. Further development and clinical validation of diagnostic tools for hvKp are warranted.

## Author contributions

Conceptualization: NW. Data curation: NW, TW, and YO. Formal analysis: NW and TM. Investigation: NW, TW, and YO. Methodology: NW and TM. Validation: NW and TM. Visualization: NW. Writing, original draft: NW. Writing, review, and editing: NW, TW, YO, and TM. All authors approved the final version of the manuscript.

During preparation of the manuscript, the authors used ChatGPT (OpenAI; GPT-5.4) to assist with English-language drafting and sentence-level editing of author-written text. The tool was not used for data analysis, data interpretation, figure generation, or drawing scientific conclusions. All output was reviewed and edited by the authors, who take full responsibility for the final content.

## Data Availability

The clinical data underlying this article are not publicly available because of privacy and ethics restrictions related to the institutional review board approval and opt-out process for this retrospective study. Additional methods and summary results are provided in the Supplementary Material.

## References

1. GBD 2019 Antimicrobial Resistance Collaborators. Global mortality associated with 33 bacterial pathogens in 2019: a systematic analysis for the Global Burden of Disease Study 2019. Lancet 2022; 400:2221–2248.

2. Russo TA, Marr CM. Hypervirulent *Klebsiella pneumoniae*. Clin Microbiol Rev 2019; 32:e00001–19.

3. Wyres KL, Lam MMC, Holt KE. Population genomics of *Klebsiella pneumoniae*. Nat Rev Microbiol 2020; 18:344–359.

4. Antimicrobial Resistance, Hypervirulent Klebsiella pneumoniae - Global situation. Available at: https://www.who.int/emergencies/disease-outbreak-news/item/2024-DON527. Accessed 3 August 2024.

5. Rapid risk assessment - Emergence of hypervirulent Klebsiella pneumoniae ST23 carrying carbapenemase genes in EU/EEA countries - first update. 2024. Available at: https://www.ecdc.europa.eu/en/publications-data/risk-assessment-emergence-hypervirulent-Klebsiella-pneumoniae-eu-eea. Accessed 13 March 2026.

6. Russo TA, Olson R, Fang C-T, et al. Identification of Biomarkers for Differentiation of Hypervirulent *Klebsiella pneumoniae* from Classical *K. pneumoniae*. J Clin Microbiol 2018; 56:e00776–18.

7. Russo TA, Alvarado CL, Davies CJ, et al. Differentiation of hypervirulent and classical *Klebsiella pneumoniae* with acquired drug resistance. MBio 2024; 15:e02867–23.

8. Russo TA, Lebreton F, McGann PT. A Step Forward in Hypervirulent *Klebsiella pneumoniae* Diagnostics. Emerg Infect Dis 2025; 31:1–3.

9. Roach DJ, Sridhar S, Oliver E, et al. Clinical and genomic characterization of a cohort of patients with *Klebsiella pneumoniae* bloodstream infection. Clin Infect Dis 2024; 78:31–39.

10. Fostervold A, Raffelsberger N, Hetland MAK, et al. Risk of death in *Klebsiella pneumoniae* bloodstream infections is associated with specific phylogenetic lineages. J Infect 2024; 88:106155.

11. Harada S, Aoki K, Yamamoto S, et al. Clinical and Molecular Characteristics of *Klebsiella pneumoniae* Isolates Causing Bloodstream Infections in Japan: Occurrence of Hypervirulent Infections in Health Care. J Clin Microbiol 2019; 57:e01206–19.

12. Barrios-Camacho H, Silva-Sánchez J, Cercas-Ayala E, et al. PCR system for the correct differentiation of the main bacterial species of the *Klebsiella pneumoniae* complex. Arch Microbiol 2021; 204:73.

13. Clinical and Laboratory Standards Institute. Performance Standards for Antimicrobial Susceptibility Testing. 34th ed. CLSI supplement M100. Clinical and Laboratory Standards Institute, 2024.

14. Magiorakos A-P, Srinivasan A, Carey RB, et al. Multidrug-resistant, extensively drug-resistant and pandrug-resistant bacteria: an international expert proposal for interim standard definitions for acquired resistance. Clin Microbiol Infect 2012; 18:268–281.

15. Charlson ME, Pompei P, Ales KL, MacKenzie CR. A new method of classifying prognostic comorbidity in longitudinal studies: development and validation. J Chronic Dis 1987; 40:373–383.

16. Wick RR, Judd LM, Gorrie CL, Holt KE. Unicycler: Resolving bacterial genome assemblies from short and long sequencing reads. PLoS Comput Biol 2017; 13:e1005595.

17. Gurevich A, Saveliev V, Vyahhi N, Tesler G. QUAST: quality assessment tool for genome assemblies. Bioinformatics 2013; 29:1072–1075.

18. Chklovski A, Parks DH, Woodcroft BJ, Tyson GW. CheckM2: a rapid, scalable and accurate tool for assessing microbial genome quality using machine learning. Nat Methods 2023; 20:1203–1212.

19. Argimón S, David S, Underwood A, et al. Rapid genomic characterization and global surveillance of *Klebsiella* using Pathogenwatch. Clin Infect Dis 2021; 73:S325–S335.

20. Lam MMC, Wick RR, Watts SC, Cerdeira LT, Wyres KL, Holt KE. A genomic surveillance framework and genotyping tool for *Klebsiella pneumoniae* and its related species complex. Nat Commun 2021; 12:4188.

21. Feldgarden M, Brover V, Gonzalez-Escalona N, et al. AMRFinderPlus and the Reference Gene Catalog facilitate examination of the genomic links among antimicrobial resistance, stress response, and virulence. Sci Rep 2021; 11:12728.

22. Seemann T. Snippy: rapid haploid variant calling and core genome alignment. GitHub. Available at: https://github.com/tseemann/snippy. Accessed 19 March 2026.

23. Croucher NJ, Page AJ, Connor TR, et al. Rapid phylogenetic analysis of large samples of recombinant bacterial whole genome sequences using Gubbins. Nucleic Acids Res 2015; 43:e15.

24. Minh BQ, Schmidt HA, Chernomor O, et al. IQ-TREE 2: New models and efficient methods for phylogenetic inference in the genomic era. Mol Biol Evol 2020; 37:1530–1534.

25. Kanda Y. Investigation of the freely available easy-to-use software “EZR” for medical statistics. Bone Marrow Transplant 2013; 48:452–458.

26. Firth D. Bias reduction of maximum likelihood estimates. Biometrika 1993; 80:27–38.

27. Kochan TJ, Nozick SH, Medernach RL, et al. Genomic surveillance for multidrug-resistant or hypervirulent *Klebsiella pneumoniae* among United States bloodstream isolates. BMC Infect Dis 2022; 22:603.

28. Tang Y, Du P, Du C, et al. Genomically defined hypervirulent *Klebsiella pneumoniae* contributed to early-onset increased mortality. Nat Commun 2025; 16:2096.

29. Yu F, Lv J, Niu S, et al. Multiplex PCR analysis for rapid detection of *Klebsiella pneumoniae* carbapenem-resistant (sequence type 258 [ST258] and ST11) and hypervirulent (ST23, ST65, ST86, and ST375) strains. J Clin Microbiol 2018; 56:e00731–18.

