## Appendix for "Abscess Complications and Prolonged Care in Five-Biomarker-Defined Hypervirulent *Klebsiella pneumoniae* Bloodstream Infection"

### Supplementary Appendix 1

Abscess Complications and Prolonged Care in Five-Biomarker-Defined Hypervirulent *Klebsiella pneumoniae* Bloodstream Infection

Naoki Watanabe, Tomohisa Watari, Yoshihito Otsuka, and Tomoh Matsumiya.

Supplementary Methods

Supplementary Table S1: Full Baseline Characteristics, Comorbidity Details, Antimicrobial Regimens, Laboratory Variables, and Outcomes by Hypervirulent Status

Supplementary Table S2: Antimicrobial Non-susceptibility by Category and Multidrug-Resistant Status

Supplementary Table S3: Missingness and Analysis Populations

Supplementary Table S4: Multivariable Logistic Regression for Abscess Complication

Supplementary Table S5: Clinical Outcomes Stratified by Number of PCR-Positive Biomarkers Among 207 *K. pneumoniae* Bloodstream Infection Episodes

Supplementary Table S6: Multivariable Linear Regression for Post-Onset Length of Stay

Supplementary Table S7: Multivariable Log-Linear Regression for Total Antibiotic Duration Among Treatment Completers

Supplementary Table S8: Firth's Penalized Logistic Regression for Prolonged Antibiotic Therapy

Supplementary Table S9: Firth's Penalized Logistic Regression for 30-Day Mortality

Supplementary Table S10: Comparison of Clinical Characteristics Between Randomly Sampled Whole-Genome-Sequenced and Non-Sequenced Classical *K. pneumoniae* Isolates

Supplementary Table S11: Sequence Type Distribution Among Sequenced hvKp, Randomly Sampled cKp, and Third-Generation Cephalosporin-Resistant Isolates

Supplementary Table S12: Capsular Locus Distribution Among Sequenced hvKp, Randomly Sampled cKp, and Third-Generation Cephalosporin-Resistant Isolates

Supplementary Table S13: Genomic Features and Abscess Complications Among Five-Biomarker-Defined Hypervirulent *K. pneumoniae* Isolates, Stratified by Capsular Locus and Sequence Type

Supplementary Note

References

#### Supplementary Methods

##### Microbiological methods

Blood cultures were processed using the BD BACTEC FX system (Becton Dickinson, Franklin Lakes, NJ). Species identification was performed using MALDI Biotyper with the MBT Compass Library version 13 (Bruker Daltonics, Bremen, Germany). Species-level differentiation within the *Klebsiella pneumoniae* species complex was performed by PCR as previously described [1].

##### PCR detection of 5 biomarkers

PCR assays for the 5 biomarkers (*rmpA, rmpA2, iucA, iroB*-PP1*, and peg-344*-PP2) were performed in 25-µL reaction mixtures containing 1× Ex Taq Buffer, 0.2 mM each dNTP, 0.4 µM of each primer, approximately 1.25 U of TaKaRa Ex Taq® Hot Start Version (Takara Bio, Shiga, Japan), and genomic DNA. Cycling conditions were an initial denaturation at 98°C for 10 s, followed by 25 cycles of 98°C for 10 s, annealing at 50°C for *rmpA* and *rmpA2*, 59°C for *iucA* and *iroB*-PP1, or 53°C for *peg-344*-PP2 for 30 s, and extension at 72°C for 30–50 s depending on amplicon size, with a final extension at 72°C for 10 min. Expected amplicon sizes were 332 bp for *rmpA*, 430 bp for *rmpA2*, 583 bp for *iucA*, 235 bp for *iroB*-PP1, and 411 bp for *peg-344*-PP2. Primer sets, annealing temperatures, and expected product sizes for *iroB*-PP1 and *peg-344*-PP2 were adopted from Russo et al [2].

##### Clinical definitions

Infection onset was classified as community-acquired if the index blood culture was collected before or within 48 hours of hospital admission in the absence of prior healthcare exposure. All other episodes were classified as healthcare-associated, including those with onset more than 48 hours after admission and those with onset within 48 hours in the presence of prior healthcare exposure. Healthcare exposure was defined as one or more of the following: outpatient chemotherapy, hemodialysis, outpatient intravenous therapy or wound care, residence in a long-term care facility, or presence of an indwelling vascular access device.

Appropriate empiric therapy was defined as initial empiric antimicrobial therapy that tested susceptible or susceptible-dose dependent against the causative isolate on antimicrobial susceptibility testing.

Post-onset length of stay was defined as days from the index blood culture collection to hospital discharge or interhospital transfer. If hospitalization continued for reasons judged unrelated to the bloodstream infection, follow-up was censored at the end of bloodstream infection-directed therapy. Episodes ending in in-hospital death were included, with length of stay calculated to death.

Total antibiotic duration was defined as the total number of days of antimicrobial therapy administered for the bloodstream infection episode and its complications, including oral therapy prescribed after hospital discharge.

##### Missing data and analysis populations

The primary outcome, abscess complication, was assessed in all included episodes. Thirty-day mortality was analyzed among episodes with available 30-day vital status. Post-onset length of stay was analyzed among episodes with available follow-up data. Total antibiotic duration was analyzed among treatment completers with available duration data; episodes ending in death before completion of therapy were excluded from the primary antibiotic-duration analysis.

##### Whole-genome sequencing

Genomic DNA was extracted using the MagLEAD instrument with the manufacturer's kit (Precision System Science, Chiba, Japan). Sequencing libraries were prepared using DNA Prep (M) Tagmentation (Illumina, San Diego, CA) and sequenced on the Illumina NextSeq X by Novogene. De novo assembly was performed using Unicycler (v0.4.8) via the Bacterial and Viral Bioinformatics Resource Center (BV-BRC; accessed November 2025–January 2026) [3,4]. Assembly quality was assessed with QUAST (v5.2.0) [5]. Assembly completeness and contamination were additionally estimated with CheckM2 (v1.1.0) using the predict workflow on de novo assemblies [6]. Predefined quality criteria required coverage depth of 100× or greater, CheckM2-estimated contamination less than 5%, total assembly length within the expected range for *K. pneumoniae* (4.5–6.5 Mb), fewer than 500 contigs, and N50 greater than 20 kb. Genomic characterization was performed using Pathogenwatch (accessed November 2025–January 2026) [7], with Klebsiella genomic typing based on Kleborate (v3.2.4) [8]. For whole-genome-sequenced isolates, *K. pneumoniae* species assignment was confirmed by genomic analysis.

Core-genome single nucleotide polymorphisms (SNPs) were identified by mapping short reads to the *K. pneumoniae* NTUH-K2044 reference genome (GenBank: AP006725.1) using Snippy (v4.6.0). A core SNP alignment was generated across all 164 isolates using snippy-core. Recombinant regions were identified and removed using Gubbins (v3.4.1) with the GTR substitution model and IQ-TREE as the tree builder [9,10]. A maximum-likelihood phylogeny was inferred from the recombination-filtered SNP alignment using IQ-TREE (v3.0.1) with the GTR+Γ4 substitution model, 1000 ultrafast bootstrap replicates, and 1000 SH-aLRT replicates. The tree was visualized and annotated using iTOL (v7.5) [11]. Annotation tracks shown in the main phylogenetic figure included study group (hvKp, intermediate [1–4 biomarkers], or cKp), selected sequence types, ESBL status, and abscess complication.

β-lactam resistance determinants were additionally re-evaluated in all 164 assemblies using AMRFinderPlus (v4.0.23) [12]. *peg-344* was additionally screened in the internal sequenced isolates by BLASTn against de novo assemblies [13]. The *peg-344* nucleotide sequence was used as the query, and isolates were considered positive when the best hit met ≥90% nucleotide identity and ≥90% query coverage (E value $<1 \times 10^{-10}$; max_target_seqs=1).

**Contig-level analysis of hvKp–ESBL convergence isolates**

Draft genome assemblies of the 3 hvKp–ESBL convergence isolates were used for contig-level analysis. Contigs carrying *bla*CTX-M genes were identified from AMRFinderPlus outputs. Reconstructed plasmid-cluster assignment and predicted mobility were then assessed using MOB-suite (v3.1.9) [14]. Hypervirulence-associated loci were summarised from existing annotation outputs and interpreted in relation to the ESBL-carrying contigs.

**Firth's penalized logistic regression**

For the supportive analysis of 30-day mortality, we used Firth's penalized logistic regression to obtain less biased estimates under sparse outcome data [15]. Given the small number of deaths in the hvKp group, we specified a parsimonious adjustment set (age and Charlson comorbidity index) to reduce overfitting and model instability in low events-per-parameter settings. Episodes with unknown 30-day vital status were treated as missing and excluded from regression models. Firth's penalized logistic regression was implemented using the R package logistf (v1.26.1).

**Length of stay: additional modelling details**

Episodes in which length of stay could not be ascertained were excluded from length-of-stay analyses. The log transformation ln(LOS+1) was used because length of stay included zero values. Regression coefficients were exponentiated to report adjusted ratios of (LOS+1) with 95% confidence intervals.

**Total antibiotic duration: additional modelling details**

Episodes with unavailable duration due to transfer, incomplete follow-up, or no treatment were treated as missing and excluded from duration analyses. To evaluate treatment burden among patients who completed antimicrobial therapy, patients who died in hospital before completion of therapy were excluded from the primary duration analysis. Sensitivity analyses repeated the adjusted models including patients who died before completion of therapy using observed duration up to death.

**Prolonged therapy threshold analyses**

Prolonged therapy was examined using prespecified thresholds (≥14, ≥21, and ≥28 days) as binary outcomes among treatment completers included in the primary antibiotic-duration analysis. A common base adjustment set was used, comprising age, sex, Charlson comorbidity index excluding age points, infection acquisition category, and primary infection source category. Because sparse outcome patterns across some infection-source strata led to separation in standard logistic regression, Firth's penalized logistic regression was used to obtain more stable and less biased estimates. In secondary threshold models, abscess complication was additionally included to assess attenuation of the association between hvKp and prolonged therapy after accounting for abscess complication.

**Biomarker stratification**

As a prespecified exploratory analysis, all 207 episodes were stratified by the number of PCR-positive biomarkers (0, 1–4, or 5 of 5). Outcomes were compared between the 0-marker and 5-marker groups using Fisher's exact test for categorical variables and the Mann–Whitney U test for continuous variables. The 1–4-marker group (n=8) was presented descriptively without formal hypothesis testing owing to the small sample size.

#### Supplementary Table

**Table S1. Full Baseline Characteristics, Comorbidity Details, Antimicrobial Regimens, Laboratory Variables, and Outcomes by Hypervirulent Status**

| Characteristic or Condition | Episodes, No. (%) | | Available (n/N) | *P value* |
|---|---|---|---|---|
| | **hvKp (n = 28)** | **cKp (n = 179)** | | |
| Comorbidities | | | | |
| Charlson comorbidity index score ≥2 | 16 (57) | 143 (80) | 207/207 | .014 |
| Chronic pulmonary disease | 2 (7) | 18 (10) | 207/207 | 1.00 |
| Congestive heart failure | 5 (18) | 37 (21) | 207/207 | 1.00 |
| Connective tissue disease | 2 (7) | 13 (7) | 207/207 | 1.00 |
| Cerebrovascular disease/ transient ischemic attack | 7 (25) | 26 (15) | 207/207 | .17 |
| Dementia | 6 (21) | 18 (10) | 207/207 | .11 |
| Diabetes (no complications) | 8 (29) | 46 (26) | 207/207 | .82 |
| Diabetes with complications | 1 (4) | 16 (9) | 207/207 | .48 |
| Hemiplegia | 2 (7) | 6 (3) | 207/207 | .30 |
| Leukemia | 0 (0.0) | 9 (5) | 207/207 | .61 |
| Mild liver disease | 1 (4) | 5 (3) | 207/207 | .59 |
| Moderate/severe liver disease | 0 (0.0) | 4 (2) | 207/207 | 1.00 |
| Lymphoma | 0 (0.0) | 23 (13) | 207/207 | .049 |
| Moderate/severe chronic kidney disease | 3 (11) | 23 (13) | 207/207 | 1.00 |
| Myocardial infarction | 4 (14) | 7 (4) | 207/207 | .045 |
| Peptic ulcer disease | 1 (4) | 24 (13) | 207/207 | .21 |
| Peripheral vascular disease | 2 (7) | 14 (8) | 207/207 | 1.00 |
| Localized solid tumor | 5 (18) | 41 (23) | 207/207 | .63 |
| Metastatic solid tumor | 3 (11) | 23 (13) | 207/207 | 1.00 |
| Immunosuppression | | | | |
| Febrile neutropenia | 1 (4) | 13 (7) | 207/207 | .70 |
| Chemotherapy | 3 (11) | 51 (28) | 207/207 | .062 |
| Systemic corticosteroids | 3 (11) | 39 (22) | 207/207 | .21 |
| Immunosuppressant use | 0 (0.0) | 9 (5) | 207/207 | .61 |
| History of transplantation | 0 (0.0) | 2 (1) | 207/207 | 1.00 |
| HIV infection | 0 (0.0) | 0 (0.0) | 207/207 | NA |
| Laboratory values | | | | |
| White blood cell count | 145 (97, 164) | 101 (50, 151) | 207/207 | .022 |
| Infection characteristics | | | | |
| Any abscess | 17 (61) | 23 (13) | 207/207 | <.001 |
| Liver abscess | 9 (32) | 12 (7) | 207/207 | <.001 |

| | | | | |
|---|---|---|---|---|
| Other intra-abdominal and pelvic abscesses | 3 (11) | 10 (6) | 207/207 | .39 |
| Lung abscess/necrotizing pneumonia | 2 (7) | 3 (2) | 207/207 | .14 |
| Urogenital/prostatic abscess | 4 (14) | 0 (0.0) | 207/207 | <.001 |
| Ocular sites | 1 (4) | 0 (0.0) | 207/207 | .14 |
| Central nervous system abscess | 2 (7) | 0 (0.0) | 207/207 | .018 |
| Musculoskeletal/soft tissue infection | 2 (7) | 0 (0.0) | 207/207 | .018 |
| Antimicrobial therapy | | | | |
| Empiric therapy (class) | | | | .69 |
| 3rd-generation cephalosporin | 6 (21) | 35 (20) | 207/207 | |
| 4th-generation cephalosporin | 1 (4) | 4 (2) | 207/207 | |
| Other cephalosporins/penicillins | 1 (4) | 9 (5) | 207/207 | |
| β-lactam/β-lactamase inhibitor | 18 (64) | 124 (69) | 207/207 | |
| Carbapenem | 2 (7) | 5 (3) | 207/207 | |
| Oral agents | 0 (0.0) | 1 (<1) | 207/207 | |
| No empiric therapy | 0 (0.0) | 1 (<1) | 207/207 | |
| Appropriate empiric therapy | 27 (96) | 151 (84) | 207/207 | .14 |
| Definitive therapy (class) | | | | <.001 |
| 3rd-generation cephalosporin | 11 (39) | 11 (6) | 207/207 | |
| 4th-generation cephalosporin | 1 (4) | 3 (2) | 207/207 | |
| Other cephalosporins/penicillins | 6 (21) | 65 (36) | 207/207 | |
| β-lactam/β-lactamase inhibitor | 5 (18) | 71 (40) | 207/207 | |
| Carbapenem | 2 (7) | 13 (7) | 207/207 | |
| Oral agents | 3 (11) | 9 (5) | 207/207 | |
| Other | 0 (0.0) | 1 (<1) | 207/207 | |
| No definitive therapy | 0 (0.0) | 6 (3) | 207/207 | |
| Outcomes | | | | |
| 14-day mortality | 3 (11) | 24 (14) | 202/207 | 1.00 |
| 30-day mortality | 4 (15) | 34 (20) | 197/207 | .61 |
| 90-day mortality | 5 (22) | 56 (35) | 183/207 | .24 |
| In-hospital mortality | 3 (11) | 34 (19) | 207/207 | .43 |

Data are n (%) or median (IQR), unless otherwise indicated. P values were calculated using Fisher's exact test for categorical variables and the Mann-Whitney U test for continuous variables and are provided for descriptive purposes. Available (n/N) indicates the number of episodes with available data out of the total in each group; percentages were calculated using available cases as the denominator. Appropriate empiric therapy was defined as initial empiric antimicrobial therapy with in vitro activity against the causative isolate.

Abbreviations: cKp, classical *Klebsiella pneumoniae*; HIV, human immunodeficiency virus; hvKp, hypervirulent *Klebsiella pneumoniae*; IQR, interquartile range; NA, not applicable.

**Table S2. Antimicrobial Non-susceptibility by Category and Multidrug-Resistant Status**

| Antimicrobial category | Agents tested | hvKp (n=28) | cKp (n=179) | *P value* |
|---|---|---|---|---|
| Penicillins | Piperacillin | 7 (25) | 66 (37) | .29 |
| Penicillins + β-lactamase inhibitors | Ampicillin–sulbactam, amoxicillin–clavulanate | 3 (11) | 47 (26) | .10 |
| Antipseudomonal penicillins + β-lactamase inhibitors | Piperacillin–tazobactam | 1 (4) | 15 (8) | .70 |
| Non-extended-spectrum cephalosporins (1st/2nd gen) | Cefazolin | 4 (14) | 42 (23) | .34 |
| Extended-spectrum cephalosporins (3rd/4th gen) | Ceftriaxone, cefepime | 3 (11) | 28 (16) | .78 |
| Cephamycins | Cefmetazole | 0 (0) | 1 (1) | 1.00 |
| Carbapenems | Imipenem, meropenem | 0 (0) | 1 (1) | 1.00 |
| Monobactams | Aztreonam | 3 (11) | 22 (12) | 1.00 |
| Aminoglycosides | Gentamicin, tobramycin, amikacin | 2 (7) | 24 (13) | .54 |
| Fluoroquinolones | Levofloxacin, ciprofloxacin | 3 (11) | 35 (20) | .43 |
| Tetracyclines | Minocycline | 0 (0) | 30 (17) | .02 |
| Folate pathway antagonists | Trimethoprim–sulfamethoxazole | 3 (11) | 46 (26) | .10 |
| Multidrug resistance | ≥3 categories | 3 (11) | 53 (30) | .04 |

Data are n (%). P values were calculated using Fisher's exact test and are provided for descriptive purposes.

Abbreviations: cKp, classical *Klebsiella pneumoniae*; hvKp, hypervirulent *Klebsiella pneumoniae*.

**Table S3. Missingness and Analysis Populations**

| Variable or outcome | Available No. | | Missing No. | | Included in main model |
|---|---|---|---|---|---|
| | hvKp (n=28) | cKp (n=179) | hvKp (n=28) | cKp (n=179) | |
| Abscess complication | 28 (100) | 179 (100) | 0 (0) | 0 (0) | Yes |
| Length of stay | 28 (100) | 178 (99) | 0 (0) | 1 (<1) | Yes |
| Total antibiotic duration | 24 (86) | 144 (80) | 4 (14) | 35 (20) | Yes, among treatment completers |
| 30-day mortality | 27 (96) | 170 (95) | 1 (4) | 9 (5) | Supportive |
| Age | 28 (100) | 179 (100) | 0 (0) | 0 (0) | Yes |
| Sex | 28 (100) | 179 (100) | 0 (0) | 0 (0) | Yes |
| Charlson comorbidity index | 28 (100) | 179 (100) | 0 (0) | 0 (0) | Yes |
| Acquisition category | 28 (100) | 179 (100) | 0 (0) | 0 (0) | Yes |
| Primary source category | 28 (100) | 179 (100) | 0 (0) | 0 (0) | Yes |

Data are n (%). For total antibiotic duration, available no refers to the primary analysis population among treatment completers. Thirty-nine episodes were excluded from this analysis, including 37 that ended in death before completion of therapy and 2 with unavailable duration data.

**Table S4. Multivariable Logistic Regression for Abscess Complication**

| Covariate | Adjusted odds ratio | 95% confidence interval | *P value* |
|---|---|---|---|
| hvKp (vs cKp) | 10.7 | 4.36–26.21 | <.001 |
| Age | 1.00 | 0.97–1.03 | .94 |
| Male sex (vs female) | 0.76 | 0.34–1.69 | .50 |
| Charlson comorbidity index | 0.91 | 0.77–1.07 | .26 |

Adjusted odds ratios are presented; odds ratios for age and Charlson comorbidity index are per 1-unit increase. Analyses used complete cases (n = 207).

Abbreviations: cKp, classical *Klebsiella pneumoniae*; hvKp, hypervirulent *Klebsiella pneumoniae*.

**Table S5. Clinical Outcomes Stratified by Number of PCR-Positive Biomarkers Among 207 *K. pneumoniae* Bloodstream Infection Episodes**

| Covariate | 0 markers (n = 171) | 1–4 markers (n = 8) | 5 markers (n = 28) | Available (n/N) | *P value*[a] |
|---|---|---|---|---|---|
| Abscess complication | 21/171 (12) | 2/8 (25) | 17/28 (61) | 207/207 | <.001 |
| Metastatic infection | 1/171 (0.6) | 0/8 (0.0) | 4/28 (14) | 207/207 | .0015 |
| 14-day mortality[b] | 23/167 (14) | 1/8 (13) | 3/27 (11) | 202/207 | 1.00 |
| 30-day mortality[b] | 33/162 (20) | 1/8 (13) | 4/27 (15) | 197/207 | .61 |
| 90-day mortality[b] | 54/153 (35) | 2/7 (29) | 5/23 (22) | 183/207 | .24 |
| Intensive care unit admission | 24/171 (14) | 1/8 (13) | 5/28 (18) | 207/207 | .57 |
| Vasopressor use | 49/171 (29) | 2/8 (25) | 7/28 (25) | 207/207 | .82 |
| Mechanical ventilation | 5/171 (3) | 0/8 (0.0) | 4/28 (14) | 207/207 | .024 |
| Length of stay, days, median (IQR) | 14 (11, 18) | 12 (10, 19) | 28 (14, 42) | 206/207 | <.001 |
| Antibiotic duration, days, median (IQR) | 14 (14, 18) | 14 (14, 15) | 43 (16, 67) | 168/207 | <.001 |

Data are n (%) or median (IQR), unless otherwise indicated. Biomarkers comprise *rmpA*, *rmpA2*, *iucA*, *iroB*, and *peg-344*, detected by PCR. All 8 isolates in the 1–4-marker group carried 3 of 5 biomarkers (*rmpA*, *iroB*, and *peg-344* positive; *iucA* and *rmpA2* negative).

[a] P values were calculated using Fisher's exact test for categorical variables and the Mann-Whitney U test for continuous variables, comparing the 0-marker and 5-marker groups, and are provided for descriptive purposes. The 1–4-marker group (n = 8) is presented descriptively without formal hypothesis testing

[b]Denominators vary because of missing vital status at 14, 30, and 90 days. Episodes with unknown vital status were excluded from the respective analyses

Abbreviations: IQR, interquartile range.

**Table S6. Multivariable Linear Regression for Post-Onset Length of Stay**

| Covariate | Model A adjusted ratio (95% CI) | Model A *P value* | Model B adjusted ratio (95% CI) | Model B *P value* |
|---|---|---|---|---|
| hvKp (vs cKp) | 1.60 (1.18–2.16) | .0026 | 1.07 (0.79–1.45) | .65 |
| Abscess complication (yes vs no) | Not applicable | Not applicable | 2.35 (1.80–3.06) | <.0001 |
| Age | 1.00 (0.99–1.00) | .23 | 1.00 (0.99–1.00) | .21 |
| Male sex (vs female) | 1.10 (0.89–1.36) | .39 | 1.13 (0.93–1.37) | .24 |
| Charlson comorbidity index | 0.99 (0.94–1.03) | .53 | 0.99 (0.95–1.03) | .73 |
| Community-acquired (vs healthcare-associated) | 1.04 (0.84–1.30) | .70 | 0.95 (0.78–1.16) | .61 |

Outcome was ln(length of stay + 1). Coefficients were exponentiated to report adjusted ratios of (length of stay + 1). Model A adjusted for age, sex, Charlson comorbidity index excluding age points, and acquisition category. Model B included all covariates in Model A plus any abscess complication. Analyses used complete cases (n = 206).

Abbreviations: CI, confidence interval; cKp, classical *Klebsiella pneumoniae*; hvKp, hypervirulent *Klebsiella pneumoniae*.

**Table S7. Multivariable Log-Linear Regression for Total Antibiotic Duration Among Treatment Completers**

| Covariate | Model A adjusted ratio (95% CI) | Model A *P value* | Model B adjusted ratio (95% CI) | Model B *P value* |
|---|---|---|---|---|
| hvKp (vs cKp) | 2.13 (1.64–2.77) | <.0001 | 1.24 (1.00–1.54) | .055 |
| Abscess complication (yes vs no) | Not applicable | Not applicable | 3.08 (2.53–3.76) | <.0001 |
| Age | 1.00 (0.99–1.00) | .54 | 1.00 (0.99–1.00) | .53 |
| Male sex (vs female) | 0.93 (0.77–1.13) | .46 | 0.97 (0.84–1.12) | .70 |
| Charlson comorbidity index | 0.99 (0.95–1.03) | .70 | 1.00 (0.97–1.04) | .76 |
| Community-acquired (vs healthcare-associated) | 1.04 (0.85–1.26) | .72 | 0.92 (0.80–1.07) | .28 |
| Primary infection source: catheter-related | 1.34 (0.71–2.53) | .37 | 1.38 (0.86–2.22) | .18 |
| Primary infection source: other | 2.09 (0.90–4.83) | .086 | 1.00 (0.53–1.89) | .99 |
| Primary infection source: respiratory | 0.91 (0.56–1.48) | .70 | 0.87 (0.60–1.24) | .43 |
| Primary infection source: unknown | 1.17 (0.88–1.56) | .29 | 1.16 (0.94–1.44) | .17 |
| Primary infection source: intra-abdominal | 1.28 (1.02–1.61) | .037 | 1.15 (0.97–1.37) | .11 |

Outcome was ln(total antibiotic duration). Coefficients were exponentiated to report adjusted ratios; ratios greater than 1 indicate longer duration. Model A adjusted for age, sex, Charlson comorbidity index, infection acquisition category, and primary infection source category. Model B included all covariates in Model A plus abscess complication. Analyses were restricted to treatment completers and used complete cases (n = 168).

Abbreviations: CI, confidence interval; cKp, classical *Klebsiella pneumoniae*; hvKp, hypervirulent *Klebsiella pneumoniae*.

**Table S8. Firth's Penalized Logistic Regression for Prolonged Antibiotic Therapy**

| Threshold | Model | Covariate | Adjusted odds ratio (95% CI) | *P value* |
|---|---|---|---|---|
| ≥14 days | Base model | hvKp | 2.21 (0.62–11.79) | .24 |
| ≥14 days | +Abs model | hvKp | 0.65 (0.14–4.15) | .62 |
| ≥14 days | +Abs model | Abscess | 23.64 (2.58–3226.46) | .0016 |
| ≥21 days | Base model | hvKp | 6.85 (2.58–19.93) | <.0001 |
| ≥21 days | +Abs model | hvKp | 1.38 (0.20–7.35) | .72 |
| ≥21 days | +Abs model | Abscess | 143.69 (25.84–2042.25) | <.0001 |
| ≥28 days | Base model | hvKp | 6.71 (2.51–19.43) | <.0001 |
| ≥28 days | +Abs model | hvKp | 1.30 (0.23–6.34) | .75 |
| ≥28 days | +Abs model | Abscess | 59.26 (15.59–354.11) | <.0001 |

Prolonged therapy was defined using prespecified thresholds of ≥14, ≥21, and ≥28 days. Base covariates were age, sex, Charlson comorbidity index, infection acquisition category, and primary infection source category. The +Abs model included all base covariates plus abscess complication. Analyses were restricted to treatment completers and used complete cases (n = 168).

Abbreviations: CI, confidence interval; hvKp, hypervirulent *Klebsiella pneumoniae*.

**Table S9. Firth's Penalized Logistic Regression for 30-Day Mortality**

| Covariate | Adjusted odds ratio (95% CI) | *P value* |
|---|---|---|
| hvKp (vs cKp) | 0.85 (0.25–2.39) | .77 |
| Age | 1.03 (1.00–1.06) | .071 |
| Charlson comorbidity index | 1.25 (1.08–1.45) | .0033 |

Adjusted odds ratios are presented; odds ratios for age and Charlson comorbidity index are per 1-unit increase. The model adjusted for age and Charlson comorbidity index and used complete cases with available 30-day vital status (n = 197).

Abbreviations: CI, confidence interval; cKp, classical *Klebsiella pneumoniae*; hvKp, hypervirulent *Klebsiella pneumoniae*.

**Table S10. Comparison of Clinical Characteristics Between Randomly Sampled Whole-Genome-Sequenced and Non-Sequenced Classical *K. pneumoniae* Isolates**

| Characteristic | WGS (n = 125) | Non-WGS (n = 54) | *P value* |
|---|---|---|---|
| Age, years, median (IQR) | 79 (70, 86) | 76 (65, 84) | .22 |
| Male sex | 74 (59) | 34 (63) | .74 |
| Charlson comorbidity index, median (IQR) | 3 (2, 5) | 3 (2, 5) | .90 |
| Onset classification | | | .13 |
| Healthcare-associated | 75 (60) | 39 (72) | .13 |
| Community-acquired | 50 (40) | 15 (28) | |
| Primary source of infection | | | .012 |
| Urinary tract | 25 (20) | 12 (22) | |
| Catheter-related | 0 (0.0) | 5 (9) | |
| Intra-abdominal | 61 (49) | 22 (41) | |
| Respiratory | 4 (3) | 1 (2) | |
| Other | 0 (0.0) | 1 (2) | |
| Unknown | 35 (28) | 13 (24) | |
| Any abscess | 15 (12) | 8 (15) | .63 |
| 30-day mortality | 22 (18) | 12 (25) | .40 |
| Length of stay, days, median (IQR) | 14 (12, 18) | 14 (10, 18) | .76 |
| Third-generation cephalosporin-resistant | 20 (16) | 8 (15) | 1.00 |

Data are n (%) or median (IQR), unless otherwise indicated. P values were calculated using Fisher's exact test for categorical variables and the Mann-Whitney U test for continuous variables and are provided for descriptive purposes. The between-group difference in primary infection source reflected catheter-related infections being present only in the non-WGS group (0/125 vs 5/54).

Abbreviations: IQR, interquartile range; WGS, whole-genome sequencing.

**Table S11. Sequence Type Distribution Among Sequenced hvKp, Randomly Sampled cKp, and Third-Generation Cephalosporin-Resistant Isolates**

| ST | hvKp (n = 28) | randomly sampled cKp (n = 125) | 3GCR (n = 31) |
|---|---|---|---|
| 23 | 10 (36) | 1 (0.8) | 0 (0.0) |
| 65 | 8 (29) | 0 (0.0) | 0 (0.0) |
| 412 | 4 (14) | 0 (0.0) | 2 (7) |
| 86 | 2 (7) | 0 (0.0) | 0 (0.0) |
| 268 | 1 (4) | 1 (0.8) | 0 (0.0) |
| 29 | 1 (4) | 0 (0.0) | 0 (0.0) |
| 893 | 1 (4) | 0 (0.0) | 1 (3) |
| 37 | 0 (0.0) | 10 (8) | 7 (23) |
| 45 | 0 (0.0) | 9 (7) | 2 (7) |
| 22 | 0 (0.0) | 5 (4) | 0 (0.0) |
| 200 | 0 (0.0) | 5 (4) | 0 (0.0) |
| 307 | 0 (0.0) | 5 (4) | 8 (26) |
| 353 | 0 (0.0) | 4 (3) | 7 (23) |
| Other | 1 (4) | 85 (68) | 4 (13) |

Data are n (%). Percentages are column percentages. Columns are not mutually exclusive; third-generation cephalosporin-resistant isolates may overlap with the hvKp and randomly sampled cKp columns.

Abbreviations: 3GCR, third-generation cephalosporin-resistant; cKp, classical *Klebsiella pneumoniae*; hvKp, hypervirulent *Klebsiella pneumoniae*; ST, sequence type.

**Table S12. Capsular Locus Distribution Among Sequenced hvKp, Randomly Sampled cKp, and Third-Generation Cephalosporin-Resistant Isolates**

| KL | hvKp (n = 28) | randomly sampled cKp (n = 125) | 3GCR (n = 31) |
|---|---|---|---|
| KL1 | 11 (39) | 2 (2) | 0 (0.0) |
| KL2 | 10 (36) | 4 (3) | 1 (3) |
| KL57 | 4 (14) | 0 (0.0) | 2 (7) |
| KL20 | 2 (7) | 1 (0.8) | 1 (3) |
| KL15 | 0 (0.0) | 9 (7) | 7 (23) |
| KL24 | 0 (0.0) | 8 (6) | 1 (3) |
| KL23 | 0 (0.0) | 7 (6) | 0 (0.0) |
| KL102 | 0 (0.0) | 6 (5) | 8 (26) |
| KL28 | 0 (0.0) | 5 (4) | 0 (0.0) |
| KL110 | 0 (0.0) | 4 (3) | 7 (23) |
| Other | 1 (4) | 79 (63) | 4 (13) |

Data are n (%). Percentages are column percentages. Columns are not mutually exclusive; third-generation cephalosporin-resistant isolates may overlap with the hvKp and randomly sampled cKp columns.

Abbreviations: 3GCR, third-generation cephalosporin-resistant; cKp, classical *Klebsiella pneumoniae*; hvKp, hypervirulent *Klebsiella pneumoniae*; KL, capsular locus.

**Table S13. Genomic Features and Abscess Complications Among Five-Biomarker-Defined Hypervirulent *K. pneumoniae* Isolates, Stratified by Capsular Locus and Sequence Type**

| Capsular locus | ST | N | KpVP-1 intact | ybt+ | Abscess |
|---|---|---|---|---|---|
| KL1 | ST23 | 10 | 10 (100) | 10 (100) | 9 (90) |
| | ST23-1LV | 1 | 1 (100) | 1 (100) | 0 (0.0) |
| KL2 | ST65 | 8 | 1 (13) | 3 (38) | 5 (63) |
| | ST86 | 2 | 2 (100) | 2 (100) | 1 (50) |
| KL20 | ST268 | 1 | 1 (100) | 1 (100) | 0 (0.0) |
| | ST893 | 1 | 1 (100) | 1 (100) | 1 (100) |
| KL54 | ST29 | 1 | 1 (100) | 1 (100) | 0 (0.0) |
| KL57 | ST412 | 4 | 2 (50) | 2 (50) | 1 (25) |

Data are n (%) unless otherwise indicated. N indicates the number of isolates. The Abscess column shows number with abscess/total (%). KpVP-1 intact was defined according to Kleborate classification.

Abbreviations: 1LV, single-locus variant; KpVP-1, *Klebsiella pneumoniae* virulence plasmid 1; ST, sequence type; ybt, yersiniabactin locus present.

#### Supplementary Note

**Contig-level genomic context of the three hvKp–ESBL convergence isolates**

Short-read assembly analysis suggested that ESBL genes and hypervirulence-associated loci were identified on separate putative plasmid-associated contigs in all three hvKp–ESBL convergence isolates, supporting independent acquisition rather than colocalization on a single reconstructed plasmid.

KML26162 (ST412/KL57; CG10014) carried *bla*CTX-M-15. The ESBL gene was identified on contig 37 (9.8 kb), which MOB-suite assigned to the putative plasmid-associated AA449/AI534 cluster; the corresponding reconstructed cluster was predicted to be conjugative. In contrast, *iuc*, *iro*, and *rmpA*/*rmpA2*-associated sequences were identified on separate contigs assigned to the distinct AA406/AI454 cluster, classified as non-mobilizable.

KML26192 (ST893/KL20; CG12072) carried *bla*CTX-M-55. The ESBL gene was identified on contig 24 (71.2 kb), assigned to the putative plasmid-associated AA277/AI090 cluster, with the corresponding reconstructed cluster predicted to be conjugative. Hypervirulence-associated sequences were again detected on separate contigs assigned to the distinct AA406/AI454 cluster, classified as non-mobilizable.

KML26241 (ST412/KL57; CG10014) carried *bla*CTX-M-15. The ESBL gene was identified on contig 39 (9.8 kb), assigned to the putative plasmid-associated AA449/AI534 cluster, with the corresponding reconstructed cluster predicted to be conjugative. As in KML26162, hypervirulence-associated sequences were detected on separate contigs assigned to the distinct AA406/AI454 cluster, classified as non-mobilizable.

The two ST412/KL57 isolates showed closely similar contig-level contexts for *bla*CTX-M-15 and hypervirulence-associated loci. Because these inferences were based on short-read draft assemblies, plasmid structures could not be resolved definitively.

#### References


1. Barrios-Camacho H, Silva-Sánchez J, Cercas-Ayala E, et al. PCR system for the correct differentiation of the main bacterial species of the *Klebsiella pneumoniae* complex. Arch Microbiol 2021; 204:73.
2. Russo TA, Olson R, Fang C-T, et al. Identification of Biomarkers for Differentiation of Hypervirulent *Klebsiella pneumoniae* from Classical *K. pneumoniae*. J Clin Microbiol 2018; 56:10.1128/JCM.00776-18.
3. Wick RR, Judd LM, Gorrie CL, Holt KE. Unicycler: Resolving bacterial genome assemblies from short and long sequencing reads. PLoS Comput Biol 2017; 13:e1005595.
4. Shukla M, Wattam AR, Aleman A, et al. BV-BRC: a unified bacterial and viral bioinformatics resource with expanded functionality and AI integration. Nucleic Acids Res 2026; 54:D715–D723.
5. Gurevich A, Saveliev V, Vyahhi N, Tesler G. QUAST: quality assessment tool for genome assemblies. Bioinformatics 2013; 29:1072–1075.
6. Chklovski A, Parks DH, Woodcroft BJ, Tyson GW. CheckM2: a rapid, scalable and accurate tool for assessing microbial genome quality using machine learning. Nat Methods 2023; 20:1203–1212.
7. Argimón S, David S, Underwood A, et al. Rapid genomic characterization and global surveillance of Klebsiella using Pathogenwatch. Clin Infect Dis 2021; 73:S325–S335.
8. Lam MMC, Wick RR, Watts SC, Cerdeira LT, Wyres KL, Holt KE. A genomic surveillance framework and genotyping tool for *Klebsiella pneumoniae* and its related species complex. Nat Commun 2021; 12:4188.
9. Croucher NJ, Page AJ, Connor TR, et al. Rapid phylogenetic analysis of large samples of recombinant bacterial whole genome sequences using Gubbins. Nucleic Acids Res 2015; 43:e15.
10. Minh BQ, Schmidt HA, Chernomor O, et al. IQ-TREE 2: New models and efficient methods for phylogenetic inference in the genomic era. Mol Biol Evol 2020; 37:1530–1534.
11. Letunic I, Bork P. Interactive Tree of Life (iTOL) v6: recent updates to the phylogenetic tree display and annotation tool. Nucleic Acids Res 2024; 52:W78–W82.
12. Feldgarden M, Brover V, Gonzalez-Escalona N, et al. AMRFinderPlus and the Reference Gene Catalog facilitate examination of the genomic links among antimicrobial resistance, stress response, and virulence. Sci Rep 2021; 11:12728.
13. Camacho C, Coulouris G, Avagyan V, et al. BLAST+: architecture and applications. BMC Bioinformatics 2009; 10:421.
14. Robertson J, Nash JHE. MOB-suite: software tools for clustering, reconstruction and typing of plasmids from draft assemblies. Microb Genom 2018; 4:10.1099/mgen.0.000206.
15. Firth D. Bias reduction of maximum likelihood estimates. Biometrika 1993; 80:27–38.
